# Integration of single cell omics with biobank data discovers *trans* effects of *SREBF1* abdominal obesity risk variants on adipocyte expression of more than 100 genes

**DOI:** 10.1101/2024.11.22.24317804

**Authors:** Mihir G. Sukhatme, Asha Kar, Uma Thanigai Arasu, Seung Hyuk T. Lee, Marcus Alvarez, Kristina M. Garske, Kyla Z. Gelev, Sandhya Rajkumar, Sankha Subhra Das, Dorota Kaminska, Ville Männistö, Hilkka Peltoniemi, Sini Heinonen, Ulla Säiläkivi, Tuure Saarinen, Anne Juuti, Kirsi H. Pietiläinen, Jussi Pihlajamäki, Minna U. Kaikkonen, Päivi Pajukanta

## Abstract

Given the fast-increasing prevalence of obesity and its comorbidities, it would be critical to improve our understanding of the cell-type level differences between the two key human adipose tissue depots, subcutaneous (SAT) and visceral adipose tissue (VAT), in their depot-specific contributions to cardiometabolic health. We integrated cell-type level RNA- and ATAC-seq data from human SAT and VAT biopsies and cell-lines to comprehensively elucidate transcriptomic, epigenetic, and genetic differences between the two fat depots. We identify cell-type marker genes for tissue specificity and functional enrichment, and show through genome-wide association study (GWAS) and partitioned polygenic risk score (PRS) enrichment analyses that the marker genes upregulated in SAT adipocytes have more prominent roles in abdominal obesity than those of VAT. We also identify *SREBF1*, a master transcription factor (TF) of fatty acid synthesis and adipogenesis, as specifically upregulated in SAT adipocytes and present in numerous SAT functional pathways. By integrating multi-omics data from an independent human cohort, we further show that the risk allele carrier status of seven abdominal obesity GWAS variants in the *cis* region of *SREBF1* affects the adipocyte expression of 146 SAT adipocyte marker genes in *trans*. We replicate this finding independently in the UK Biobank by showing that the partitioned abdominal obesity PRSs of the *trans* gene sets differ by the regional *SREBF1* risk allele carrier status. In summary, we discover the master TF, *SREBF1*, driving the SAT adipocyte expression profiles of more than a hundred of adipocyte marker genes in *trans*, a finding that indicates that human *trans* genes can be identified by integrating single cell omics with biobank data.

## Introduction

Subcutaneous (SAT) and visceral adipose tissue (VAT) are the two key fat depots in humans. SAT has many important functional roles, including lipogenesis, lipolysis, hormonal, and endocrine functions, and SAT is also the fat depot that expands most in the presence of obesity^1,2^. Efficient adipogenesis (i.e. differentiation of preadipocytes to adipocytes) is critical for this expansion and buffering against lipotoxicity and low-grade inflammation, the hallmark of obesity^1,3^. VAT, i.e. the deeper intra-abdominal fat that lines internal organs, has a lesser capacity to expand in the presence of obesity and is even more prone to pro-inflammatory profiles than SAT^1,4,5^. It has been postulated that efficient SAT adipogenesis is likely relevant for this VAT inflammation as well^6^; however, the actual tissue- and cell-type-specific functions of VAT are not comprehensively understood in humans^4^, likely partly reflecting the practical difficulties in obtaining human VAT samples. Thus, it would be crucial to advance our understanding of the cell-type level transcriptional differences between these two human main fat depots and how they relate to depot-specific contributions to cardiometabolic health and disease.

Previous bulk tissue RNA-seq studies have successfully identified *cis* regulatory variants and their targets genes in local *cis* expression quantitative trait locus (*cis*-eQTL) analyses^7–10^ Furthermore, SAT bulk tissue analysis have detected abdominal obesity associated co-expression networks, regulated by transcription factors (TFs), such as *TBX15*^11^, suggesting that TFs regulate cardiometabolic trait -associated gene expression in fat depots in *trans*. However, identification of *trans* regulatory variants and their target genes has been challenging in SAT and VAT due to the extensive multiple testing issue of the *trans*-eQTL analysis and small samples sizes of SAT and VAT data available for study, which has hampered powerful enough *trans*-eQTL discovery. Accordingly, very few *trans* signals have been identified^12,13^ and replicated in SAT or VAT. Thus, it is not well understood how variant level differences in TFs, especially in disease-connected GWAS effect alleles, contribute to downstream target genes in *trans* in bulk tissues, and even less is known about single cell level trans regulatory variants their target genes in SAT or VAT.

To advance *trans* gene discovery, we first performed dual-tissue single nucleus RNA-sequencing (snRNA-seq) from the same individuals’ SAT and VAT biopsies, which identified the unique cell-type marker genes in each fat depot for the main cell-types between SAT and VAT. We then separated these markers into those specific to SAT, specific to VAT, and shared between the two tissues. This design helped us identify a key adipose tissue TF, Sterol Regulatory Element Binding Transcription Factor 1 (*SREBF1*)^14,15^, among the unique SAT marker genes, present in 87% of functional pathways of SAT adipocytes, suggesting that it exhibits an important *trans* regulatory role. Next, we found regional abdominal obesity -associated GWAS variants landing in SAT adipocyte open chromatin in the *cis* region of *SREBF1*. Using a large set of SAT snRNA-seq data from an independent human cohort, we then identify more than one hundred *SREBF1* target genes, the adipocyte expression of which differ by the *SREBF1* abdominal obesity GWAS risk variant carrier status. Finally, we discover by building regional PRSs for the *SREBF1 trans* target genes in the independent UK Biobank, that their abdominal obesity PRSs differ by the risk allele carrier status of the *SREBF1* abdominal obesity GWAS variants, providing thus converging evidence to the single cell level data about the *trans* effects of this key TF. Overall, our study integrate single cell omics and biobank data to identify a master SAT TF and a large number of its *trans* regulated adipocyte genes for abdominal obesity.

## Methods

### Study cohorts

#### KOBS cohort

In our dual tissue study design, we analyzed both subcutaneous (SAT) and visceral adipose tissue (VAT) single nucleus RNA-sequencing (snRNA-seq) data from matching seven Finns with obesity (2 females and 5 males) from the Kuopio Obesity Surgery Study (KOBS) cohort^16,17^. This cohort comprises individuals with severe obesity (BMI>30 kg/m^2^) who underwent bariatric surgery, and whose SAT and VAT biopsies were both collected at the surgery baseline for these single-cell level omics analyses. The mean BMI and age of these seven individuals are 38.03 kg/m^2^ (SD=1.76) and 54.11 years (SD=2.03). All participants provided informed written consent. The study was approved by the local ethics committee. All research conformed to the principles of the Helsinki Declaration.

#### UKB cohort

For our genome-wide association study (GWAS) enrichment, polygenic risk score (PRS), and partitioned heritability analysis, we used the UK Biobank (UKB)^18^. Given that our single-cell level data are of European origin, we only included the unrelated European-origin UKB participants (n=391,701) in our analysis. As described previously^18,19^, the genotype data for these individuals were generated using two highly overlapping genotype arrays, Applied Biosystems UK BiLEVE Axiom Array (807,411 markers) and Applied Biosystems UK Biobank Axiom Array (825,927 markers)^18^, and imputed using the Haplotype Reference Consortium (HRC), UK10K, and 1000 Genomes panels^19^. Data from UKB were accessed under application 33934.

#### RYSA cohort

Finnish individuals with obesity were recruited for the RYSA bariatric surgery study at the Helsinki University Hospital, Helsinki, Finland, as described previously^20^. In this study, we used the operation time-point SAT snRNA-seq data from 68 RYSA participants with obesity to test whether adipocyte expression of SAT adipocyte marker genes is affected in *trans* by the risk allele carrier status of seven WHRadjBMI GWAS variants in the *SREBF1 cis* region. The RYSA study was approved by the Helsinki University Hospital Ethics Committee, and all participants provided a written informed consent. All research conformed to the principles of the Helsinki Declaration.

### Genotype quality control and imputation in the KOBS and RYSA cohorts

We genotyped DNAs of KOBS participants using the Infinium Global Screening Array-24 v1 (Illumina). We performed quality control (QC) on the genotype data using PLINK v1.9^21^ by excluding individuals with missingness >2% and removing unmapped, strand ambiguous, and monomorphic SNPs in addition to variants with missingness >2% and Hardy-Weinberg Equilibrium (HWE) *p*-value<10^-6^. We further imputed biological sex using the ‘--sex-check’ function in PLINK v1.9^21^ and cross-checked with the reported sex of each individual.

We performed genotype imputation against the HRC reference panel version r1.1 2016^22^ on the Michigan imputation server after removing duplicate variants and variants with allele mismatch with the reference panel. Strand flips or allele switches needed to match the reference panel were performed on the server before haplotype phasing using Eagle v2.4^23^ and genotype imputation using minimac4^24^. We performed QC on the imputed genotype data by removing SNPs with imputation score R^2^<0.3 and HWE *p*-value<10^-6^ for the downstream analyses.

We genotyped the RYSA participants’ DNAs using the Infinium Global Screening Array-24 v1 (Illumina). Genotype data QC, imputation, and post-imputation QC were performed for the RYSA cohort as described above.

### Nuclei isolation in VAT biopsies from the KOBS cohort

To isolate nuclei from the snap-frozen VAT biopsies from the KOBS participants, we first combined and minced the VAT samples in a petri dish over dry ice and immediately transferred the minced tissue into a 500 μl chilled 0.1X Lysis Buffer consisting of 10 mM Tris-HCl, 10 mM NaCl, 3 mM MgCl_2_, 0.1% Tween-20, 0.1% IGEPAL CA-630, 0.01% Digitonin, 1% BSA, 1 mM DTT, and 1 U/μL RNase inhibitor. After a 15-minute incubation period, the lysate mixed with a 500 μl chilled Wash Buffer containing 10 mM Tris-HCl, 10 mM NaCl, 3 mM MgCl_2_, 1% BSA, 0.1% Tween-20, 1 mM DTT, and 1 U/μL RNase inhibitor was passed through a 70 μm Flowimi Cell Strainer into a 2 ml tube. Nuclei were centrifuged at 500 rcf for 5 minutes at 4°C and the supernatant was removed without disrupting the nuclei pellet, followed by a resuspension in a 1 ml chilled Wash Buffer. Nuclei were passed through a 40 μm Flowimi Cell Strainer into a 2 ml tube and centrifuged at 500 rcf for 5 minutes at 4°C. We removed the supernatant without disrupting the nuclei pellet and resuspended in a 30 μl chilled Diluted Nuclei Buffer containing 1X Nuclei Buffer (10x Genomics), 1 mM DTT, and 1 U/μl RNase inhibitor. We measured the concentration and overall quality of the nuclei using Countess II FL Automated Cell Counter after staining with trypan and DAPI and used the Single Cell Multiome ATAC + Gene Expression Reagent Kit (10x Genomics) for the joint snRNA- and snATAC-seq library construction. We analyzed the quality of cDNA and libraries using Agilent Bioanalyzer and sequenced the libraries on an Illumina NovaSeq X Plus (snRNA) and Next seq 500 (snATAC) with a target sequencing depth of 400 million read pairs for both snRNA- and snATAC-seq.

### Nuclei isolation in SAT biopsies of the KOBS and RYSA cohorts

We performed SAT snRNA-seq experiments on the snap-frozen SAT biopsies from the KOBS participants, as previously described^11^. Briefly, we first pooled approximately 100 mg of each biopsy and isolated nuclei from the pooled biopsies as described earlier^11^. Next, we measured the concentration and overall quality of the nuclei using Countess II FL Automated Cell Counter after staining with trypan and DAPI and used the Single Cell 3’ Reagent Kit v3.1 (10x Genomics) for the library construction. We analyzed the quality of cDNA and gene expression library using Agilent Bioanalyzer and sequenced the library on an Illumina NovaSeq SP with a target sequencing depth of 600 million read pairs.

In this study, we used the operation time-point SAT snRNA-seq data from 68 participants with obesity of the RYSA cohort, generated as part of the full RYSA cohort, by randomly pooling 8 SAT samples/batch for nuclei isolation. We isolated nuclei and constructed libraries for SAT snRNA-seq from each batch as described above and sequenced libraries from all batches together on an Illumina NovaSeq S4 with a target sequencing depth of 400 million read pairs per batch.

### Processing of the SAT snRNA-seq data from the RYSA cohort

We aligned the raw snRNA-seq data in a FASTQ format file for each batch against the GRCh38 human genome reference and GENCODE v42 annotations^25^ using STAR v2.7.10b^26^ with the ‘-- soloFeatures GeneFull’ option to account for full pre-mRNA transcripts. We evaluated the quality of the raw and mapped snRNA-seq data using FastQC. Next, we used DIEM v2.4.0^27^ to remove empty droplets and droplets with high amounts of ambient RNA. We set droplets with UMI<500 as debris, used *k*=50 for the initialization step with k-means clustering, and otherwise used the default parameters. After the clustering step, we removed clusters of highly contaminated droplets characterized by low average UMIs, low average number of unique genes detected (nFeatures), high percentage of reads mapped to the mitochondrial genome (%mito), and high number of mitochondrial and ribosomal genes as top expressed features. Next, we used Seurat v4.3.0.1^28^ to remove low-quality droplets with UMI<500, nFeatures<200, %mito>10%, and spliced read fraction≥75%, log-normalize gene counts using the ‘NormalizeData’ function, identify top 2,000 variable genes excluding mitochondrial and ribosomal genes using the ‘FindVariableFeatures’ function, scale the normalized gene counts to mean 0 and variance 1 using the ‘ScaleData’ function, perform principal component analysis (PCA) using the ‘RunPCA’ function, and cluster the nuclei with a standard Louvain algorithm, first 30 PCs, and a resolution of 0.5. For the remaining nuclei, we removed contaminated counts using DecontX^29^ with the previously removed low-quality nuclei as the background and the Seurat cluster assignment as the ‘z’ and removed additional low-quality nuclei with UMI<500, UMI>30,000, nFeatures<200, and %mito>10% based on the remaining clean counts. To identify the originating participant of each nucleus, we used demuxlet from popscle software tool^30^ with ‘-- min-MQ 30’ and high-quality imputed genotype data. We only included nuclei classified as a singlet and assigned the best matching participant to identify the originating participant for each nucleus. Next, we used DoubletFinder^31^ to identify and remove any remaining doublets. As DoubletFinder requires predicted number of doublets in each dataset, we performed pN-pK parameter sweeps on a subset of 10,000 nuclei to select pN of 0.25 and the most optimal pK value maximizing mean-variance normalized bimodality coefficient for each batch, as previously recommended^27^.

After performing the QC, we used Seurat v4.3.0.1^28^ to merge all remaining high-quality droplets from the batches and subset for nuclei originating from operation time-point samples of the 68 participants from the RYSA cohort included in this study. In the subset, we kept only the genes with at least 3 raw counts in at least 3 nuclei^29^ and performed gene count normalization, variable gene identification, data scaling, and PCA, as described above. To account for variation in gene expression driven by batch effect, we used Harmony v1.0.3^32^ to integrate on batch and clustered the nuclei with a standard Louvain algorithm, first 30 reductions from Harmony, and a resolution of 0.5. Cell-type annotations were performed using SingleR v1.8.1^33^, as described above, with the SAT single-cell and snRNA-seq data from the previously published adipose tissue atlas by Emont et al. as a reference^34^.

### Processing of the SAT VAT snRNA-seq data from the KOBS cohort

We aligned the VAT and SAT snRNA-seq data from the seven KOBS biopsies to the GRCh38 human genome reference with Ensembl annotations using CellRanger-arc -count v2.0.0^35^ and CellRanger -count v6.1.1^35^ for VAT and SAT, respectively. As the VAT sequencing from the seven KOBS VAT biopsies was generated using the 10x Genomics Multiome platform, we simultaneously profiled RNA and ATAC in each cell. We subset the data to include the nuclei passing the QC for both RNA and ATAC, as recommended by CellRanger-arc, while only including the VAT snRNA-seq data in the downstream analyses of this study. The KOBS SAT snRNA-seq data were generated as a single modality and processed using the same reference (GRCh38 human genome) but with the base software for CellRanger. Similarly to VAT, we only included the nuclei from SAT that passed the recommended filtering from CellRanger. For the remaining SAT and VAT nuclei, we separately further filtered out those containing ambient reads using DIEM^27^ through the removal of droplets from a debris cluster that reflected high amounts of background RNA or low levels of nuclear RNA. To remove genetic doublets and demultiplex the KOBS VAT Multiome samples and SAT snRNA-seq samples back to their individuals of origin, we ran demuxlet^30^ against the imputed genotype data of the individuals. We removed all SAT and VAT cells that were not classified as singlets. We then ran DecontX^29^ to remove contaminated reads within each droplet and filtered out nuclei with high levels of ambient RNA, keeping those with UMIs over 200 and mitochondrial read percentage<10%. These QC processes resulted in 3,216 nuclei for VAT and 3,516 nuclei for SAT from the same individuals.

### Integration and clustering of SAT and VAT snRNA-seq data from the KOBS cohort

To find differences between SAT and VAT cell-type level gene expression from the KOBS biopsy samples, we performed integration between the tissues using Seurat v4.3.0^28^. The count data for nuclei from each tissue were first log-normalized using the NormalizeData function of Seurat^28^, with the default scaling factor of 10,000. Using these normalized count data, the top 2000 variable genes per tissue were calculated using the FindVariableFeatures function. Afterwards, we integrated the two tissues using canonical correlation analysis (CCA) with the IntegrateData function of Seurat^28^. Following integration, the normalized read counts were scaled with a mean of 0 and variance of 1. In the integrated samples with a total of 6,732 cells from across the SAT and VAT snRNA-seq data, we performed PCA and identified clusters using 30 PCs and a resolution of 0.8.

### SnRNA-seq cell-type assignment and unique marker gene identification in the KOBS SAT and VAT biopsy samples

We used SingleR v2.0.0^33^ with a previously published human single-cell adipose atlas^34^ as a reference to annotate the data at the individual cell level^36^. We ran the FindAllMarkers function in Seurat^28^ using *only.pos* = TRUE and min.cells.group=50 to identify marker genes of each cell-type. We only included cell-types that consisted of at least 50 nuclei from the integrated tissues. P-values were adjusted for multiple testing using the Bonferroni approach. To identify marker genes unique per cell-type, we removed genes detected as marker genes (padj<0.05) for multiple cell-types. These marker genes are shown in Supplementary Table 1.

### Tissue-specific marker gene identification

To find differences in the expression profiles of the three most prevalent SAT and VAT cell-types (adipocytes, macrophages, and adipose stem and progenitor cells (ASPC)) in the KOBS cohort, we identified three separate sets of cell-type marker genes. These three sets comprised cell-type marker genes specific to SAT (MGSS), VAT (MGSV), and shared between the tissues (MGSBT). We identified tissue-specific markers using the FindMarkers function of Seurat^28^, using a log_2_ fold change threshold of ≥0, to compare the SAT and VAT data of each cell-type. In this tissue-specific marker gene identification, we only included the genes identified as unique markers for each cell-type above. We adjusted p-values for multiple testing using FDR<0.05 for the total number of cell-type-specific markers from above. Using this adjustment method, we identified genes that are not only cell-type-specific but also show differential expression (DE) between VAT (MGSV) and SAT (MGSS). We also identified a third group of marker genes for each cell-type, i.e., those that showed no significant DE (FDR>0.05) between SAT and VAT. We classified these as marker genes shared between the SAT and VAT tissues (MGSBT). Overall, this resulted in 9 cell-type marker gene sets, i.e., three per cell-type (macrophages, adipocytes, and ASPC) with no overlap between them. These 9 sets are listed in Supplementary Table 2.

### Identification of enriched functional pathways

After identifying the 9 sets of cell-type marker genes, as described above, we ran WebGestalt^37^ to functionally characterize each set, using all genes with non-zero expression in at least two cells in the cell-type^34^ irrespective of the tissue origin, as the background. For each set, we tested for significant (FDR<0.05) overrepresentation of genes from Gene Ontology (GO) biological processes, cellular components, and molecular functions. We then identified the top pathway genes (i.e. the genes that appeared in >1 significantly enriched pathways) by ranking each gene by the number of times it appeared in the identified pathways.

### GWAS enrichment analysis of the cell-type marker genes

We assessed the 9 sets of tissue-specific and tissue-shared cell-type marker genes for significant enrichments of genetic associations with BMI, WHRadjBMI, T2D, and MASLD using MAGENTA v2.4 (Meta-Analysis Gene-set Enrichment of variaNT Associations)^38^.

We first downloaded publicly available European-only GWAS summary statistics for T2D^39^, while for WHRadjBMI and BMI, we used the UKB GWAS data we generated, as described above. For every outcome, we then used the summary statistics to generate association scores for each gene within the gene set based on the p-values of all variants within a 500kb upstream and downstream window. Gene score-cutoff tests using the 75th percentiles of the scores of all genes as cutoffs were performed to evaluate enrichment. While MAGENTA^38^ filters out genes near one another, we retained all genes per set in our analyses due to our small gene set sizes.

### Construction of marker gene -based regional and genome-wide PRSs for abdominal obesity, BMI, and type 2 diabetes in UKB

We built both marker gene -based regional and genome-wide PRSs for waist-hip ratio adjusted for BMI (WHRadjBMI), body mass index (BMI), and type 2 diabetes (T2D) for all unrelated Europeans from UKB^18^. In our analysis, WHRadjBMI was used as a well-established proxy for abdominal obesity^11^. The genome-wide PRSs for these traits were constructed using all SNPs (see below), whereas the regional PRSs for each trait were created using the *cis* regional SNPs (gene+/-500kb) of each of the autosomal marker gene sets separately (MGSBT, MGSS, and MGSV) for the three main cell-types (adipocytes, macrophages, and ASPCs). We excluded the set that contained shared SAT/VAT macrophage marker genes as it comprised only 9 genes, and, therefore, it remained too small for reliable regional PRS building given the technical issues with overfitting in PRS^40^. As WHRadjBMI is a highly sex-specific trait^11^, we also built the regional WHRadjBMI PRSs for males (n= 88,988) and females (n=104,614) separately. Thus, in addition to the genome-wide PRSs, we constructed 24 regional PRSs for WHRadjBMI and 8 regional PRSs for T2D and BMI, respectively.

### Construction of WHRadjBMI and BMI PRSs

To construct the WHRadjBMI and BMI PRSs, we randomly selected 50% of the unrelated Europeans from UKB (n=195,863) to generate GWAS summary statistics as the base data for the PRS, and applied the PRS model in the remaining 50% of the cohort (i.e., the target group). For the GWASs, the data were filtered to only retain variants with MAF >1% and INFO>0.8. We also removed individuals with >1% of their genotypes missing or extreme heterozygosity, as well as variants missing in >1% of the subjects or violating Hardy-Weinberg equilibrium from the genotype data target group^41^. We conducted the GWASs of WHRadjBMI and BMI using linear-mixed model approach of BOLT-LMM v2.3.6^23^, with age, age^2^, sex, the top 20 genetic PCs, testing center, and genotyping array as covariates, where we applied a rank-based inverse normal transformation for each outcome. For WHRadjBMI, given the sex-specificity of the outcome^11,42,43^, we normalized twice, first in all individuals, and then for males and females separately. Next, we fit the PRS models and computed the PRSs for WHRadjBMI and BMI for the individuals in the target group using the split-validation mode from lassosum^44^ (n=193,602) and the filtered GWAS summary statistics as the base data for the PRS models.

### Construction of T2D PRSs

To construct the T2D PRSs, we used the publicly available T2D GWAS summary statistics of Europeans without UKB from the DIAGRAM Consortium^45^ as the base data. We built the PRS models using the target group genotype data employing PRScs^46^, with the provided LD matrix from the Europeans in the 1000 Genomes Project as the LD reference, and applied the models to the target group using PLINK.

### Calculating the explained variance in the trait by the PRSs

We calculated the incremental variance explained (R^2^) in the trait against a null model containing the covariates of age, age^2^, the top 20 genetic PCs, testing center, genotyping array, and sex using lassosum^44^. For T2D, we calculated a delta AUC, similarly to the incremental variance from above.

We evaluated the significance of each regional PRS by ranking the observed incremental R^2^ or delta AUC against the incremental R^2^/delta AUC of 10,000 PRSs similarly built from the *cis* regional variants of random gene sets of the same size as in the regional PRS set, using all expressed genes (counts>=1 in at least 2 nuclei)^34^ in the cell-type of interest as a background. The P-value was defined as the number of permutations in which the R^2^/delta AUC is larger than the calculated PRS divided by 10,000.

### Partitioned heritability assessment of PRS-enriched gene sets

We performed a partitioned heritability analysis with LD Score regression (LDSC)^47,48^ to assess the genes with WHRadjBMI R^2^ enriched PRSs for enrichment in the WHRadjBMI heritability relative to the genome, similarly as in Finucane et al.^48^. Briefly, we used LDSC to estimate the LD scores from all SNPs in the genome, as well as from all SNPs residing with the *cis* regions of each PRS-enriched gene set. Scores were constructed in 76,758 randomly selected, unrelated Brits from UKB (35,257 males, 41,501 females) for computational efficiency, and as in Finucane et al.^48^, we only included SNPs with MAF>5% for the analysis. We then used the same WHRadjBMI GWAS summary statistics as in the PRS analysis to compute the WHRadjBMI heritability of the genome and marker gene annotations and test each marker annotation for significant (p<0.05) heritability enrichment, defined as the proportion of heritability divided by the proportion of the genome from the annotation^48^.

### Longitudinal RNA- and ATAC-sequencing during human SAT primary preadipocyte differentiation

We previously performed a longitudinal adipogenesis experiment using human SAT primary preadipocytes and generated longitudinal bulk RNA- and ATAC-seq data on samples collected at the 0 day (0d), 1d, 2d, 4d, 7d, and 14d timepoints, with 4 isogenic replicates per timepoint, as described earlier^7,49^. Peaks from the ATAC-seq were filtered to remove blacklisted regions and identify consensus peaks, as described in detail previously^7^.

### Differential expression (DE) analysis across six adipogenesis time points

We examined the longitudinal expression patterns of the top pathway-enriched genes from the adipocyte MGSS set (n=43 genes) using ImpulseDE2 v0.99.10^50^. We used the runImpulseDE2 function with default parameters and *boolCaseCtrl*=FALSE, *boolIdentifyTransients*=TRUE, and *scaNProc*=1. P-values were corrected for multiple testing using FDR<0.05. As the adipogenesis experiment was conducted in SAT preadipocytes, we only ran this DE analysis for genes in the adipocyte MGSS set.

### Identification of longitudinally co-expressed clusters of adipocyte pathway genes and their regulators during human adipogenesis

To search for longitudinal co-expression patterns among the adipocyte pathway genes across human adipogenesis, we ran DPGP v0.1^51^ to cluster genes by their expression trajectories. We ran DPGP with the timepoint data of the adipogenesis experiment described above and only included the genes that we identified as significantly longitudinally DE (FDR<0.05) during human adipogenesis from ImpulseDE2^50^ and expressed across adipogenesis (n=42).

### Identification of co-accessible peaks during human adipogenesis within the *cis* region of *SREBF1*

To identify co-accessible peaks in the *SREBF1 cis* region during human adipogenesis, we used the longitudinal ATAC-seq data from the human SAT primary preadipocyte differentiation experiment described above. We first identified SAT adipocyte peaks within ±500 kb of *SREBF1*, a key SAT adipocyte marker gene. To determine temporal differential accessibility (DA) of these SAT peaks and identify clusters of longitudinally co-accessible peaks, we ran ImpulseDE2^50^ as described above on the accessibility counts for the peaks, subset the data to the peaks passing an FDR threshold<0.05, and then clustered the DA peaks (n=120) using DPGP^51^, as described above. We only included the clustered peaks that passed a cluster assignment threshold of probability>0.9.

### Identification of WHRadjBMI GWAS variants within the longitudinally co-accessible SAT adipocyte peaks in the *SREBF1 cis* region

We next investigated the 5 longitudinally co-accessible SAT adipocyte peaks discovered in the SAT adipogenesis experiment within the *cis* regions of *SREBF1* for WHRadjBMI GWAS variants. Accordingly, we overlapped the positions of the genome-wide significant (p<5× 10^-8^) WHRadjBMI GWAS variants using the summary statistics from a previously published extensive WHRadjBMI GWAS of the GIANT–UKB meta-analysis^52^ with the positions of the co-accessible SAT adipocyte peaks.

### Identification of adipocyte MGSS genes by risk allele status of the WHRadjBMI GWAS variants in the *SREBF1* region

We tested whether the risk allele carrier status of the seven WHRadjBMI GWAS variants residing in the longitudinally co-accessible SAT peaks in the *SREBF1* region affects the SAT adipocyte expression of *SREBF1* and other SAT adipocyte markers genes in *trans* using RYSA SAT adipocyte snRNA-seq data. Briefly, for each variant, we labeled each individual from the RYSA cohort with zero copies of the GWAS trait-increasing allele as “non-carriers” and those with one or two copies of the trait-increasing allele as “carriers,” and then labelled each individual cell in the cohort as coming from a “carrier” or a “non-carrier” individual. We subset the SAT snRNA-seq data to adipocytes and ran the FindMarkers function in Seurat^28^ with logfc.threshold = 0 between cells labeled “carriers” and “non-carriers” while only testing the adipocyte MGSS gene set and adjusting the p-values for multiple testing using FDR<0.1. We repeated this for each of the seven SNPs. For each *trans* gene set, we built module scores using the AddModuleScore function in Seurat v4.3.0^28^ in SAT adipocytes from the RYSA cohort to determine the average expression across all adipocytes between carriers and non-carriers.

### Identification of adipogenesis open chromatin peaks by cell-type

To identify upregulated and shared peaks at the preadipocyte and adipocyte stages of human adipogenesis, we ran DESeq2^53^ on the raw peak count data generated using ATAC-seq at the day 0 and 14 time points in the SAT adipogenesis experiment described above. Peaks with a log_2_FC>0.1 and Bonferroni adjusted P (padj)<0.05 were classified as upregulated in preadipocytes, while the peaks with a log_2_FC<-0.1 and padj<0.05 were classified as upregulated in adipocytes. The remaining peaks were classified as shared between the human preadipocytes and adipocytes.

### Construction of abdominal obesity PRSs in UKB for the adipocyte marker genes affected by the seven regional *SREBF1* variants in *trans*

To investigate the abdominal obesity risk of the regional variants in the *trans* gene sets, the adipocyte expression of which is up- or downregulated by the *SREBF1* variant carrier status, we constructed partitioned PRSs using the *cis* regional SNPs of these *trans* gene sets. The PRSs were built separately for each up/downregulated *trans* gene set for the seven SNPs, resulting in 14 total unique PRS sets. As abdominal obesity risk differs by sex^42,43^, we analyzed these partitioned abdominal obesity PRSs in all individuals, females, and males separately. Due to the small region sizes and to allow for fine-grained control of the SNPs comprising each PRS, we used a clumping and thresholding (C+T) approach with PLINK v1.9 for the PRS construction, with the same GWAS summary statistics for WHRadjBMI described above, as the base data. We partitioned the remaining 50%, not used for the GWAS, into two cohorts, a 30% test group (n=115,120) for performing the LD clumping and learning the optimal thresholding parameter, and a 20% validation group (n=76,758) to apply the learned PRS model and perform downstream analyses. We performed QC on the test and validation genotype data, similarly as described above.

To build the partitioned abdominal obesity PRSs, we first performed LD clumping on the full genotype data of the test groups with PLINK v1.9 using R^2^ of 0.2 and window size of 250 kb, and built the PRSs from the independent SNPs passings p-value thresholds ranging from 5×10^-8^ to 0.5 to empirically determine the p-value threshold that maximizes the incremental variance explained in WHRadjBMI in the test group. Next, for each *trans* gene set, we identified all C+T SNPs that landed in the adipocyte upregulated open chromatin peaks (described earlier), using the empirically determined p-value thresholds of p<0.05, p<0.005, and p<0.1 for all individuals, females, and males, respectively. To avoid direct *SREBF1 cis* effects on the abdominal obesity risk, we removed the seven SNPs in the *SREBF1* region as well as any C+T SNPs in LD (R^2^>0.1) with any of these seven SNPs from these partitioned PRS analyses. We computed PRSs for the sets that contained >= 5 valid SNPs. As explained before, the incremental variance (R^2^) was calculated for each PRS and compared against 10,000 permuted PRSs built from the same numbers of C+T SNPs.

After computing individual level WHRadjBMI PRSs, we compared the polygenic risk, i.e., the PRS magnitudes, in the regional PRSs of the *trans* gene sets between the *SREBF1* variant carriers and non-carriers. First, we classified individuals in the UKB PRS cohort into “carriers” or “non-carriers” in the same way as in the RYSA cohort. Next, we performed a two-sided Wilcoxon test between the WHRadjBMI PRSs of the carriers and non-carriers within each group while correcting for multiple testing using Bonferroni.

## Results

### Study design

The summary of the study design is shown in Supplementary Figure 1. Briefly, by first identifying cell- and tissue-type expression profiles that drive functional differences between the two key human fat depots, subcutaneous adipose tissue (SAT) and visceral adipose tissue (VAT) and genetic enrichments of these SAT and VAT -specific genes for abdominal obesity, we discovered a transcription factor (TF) gene preferentially involved in numerous functional SAT pathways over VAT. We then further investigated the cell-type level regulatory mechanisms of this TF, and discovered that the regional abdominal obesity GWAS variants of this TF affect the adipocyte expression of tens of SAT adipocyte marker genes in *trans* in the independent RYSA cohort. Finally, we confirmed the identified *trans* effects of the TF variants by comparing the partitioned regional PRSs of the *trans* genes by their risk allele carrier status at the biobank level (Figure 1a).

**Figure 1.**
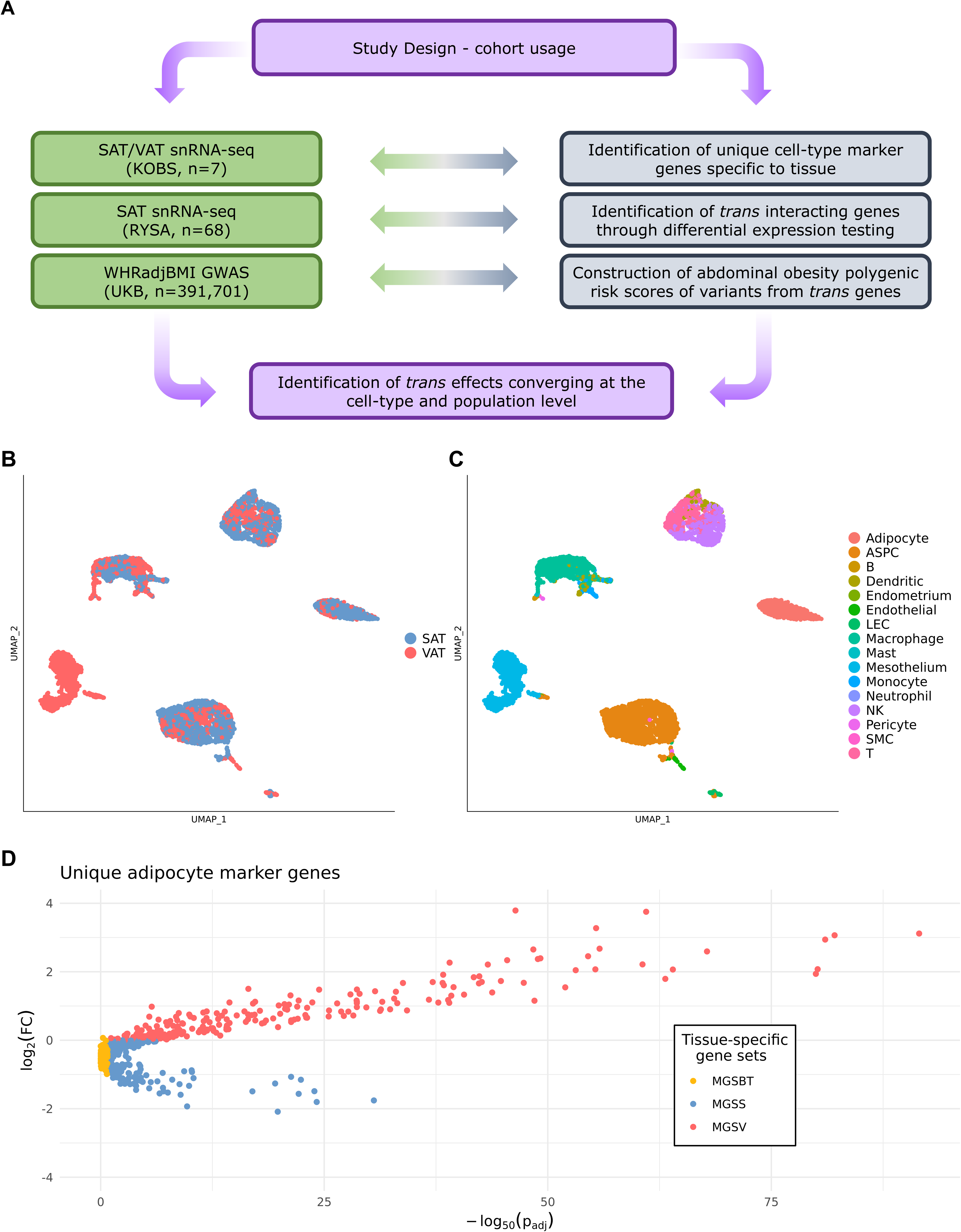
Adipose single nucleus RNA-sequence data from the KOBS participants’ subcutaneous adipose tissue (SAT) and visceral adipose tissue (VAT) identify distinct sets of tissue-specific and shared cell-type marker genes. (a) The study design visualized through cohort usage. The data from the individuals in the KOBS cohort were used to identify cell-type marker gene sets, while the data from the individuals in the RYSA cohort were used to identify *trans* gene sets by the carrier status of the *SREBF1* risk alleles. Lastly, all polygenic risk scores were built using individuals from the UK Biobank. (b-c) The integrated SAT and VAT single nucleus RNA-seq data (n=6,732 nuclei) from 7 KOBS participants are visualized in a Uniform Manifold Approximation and Projection (UMAP) and colored by (b) fat depot of origin and (c) cluster cell-type identity, while mapping each cluster to one of the 16 identified cell-types. ASPC indicates adipose stem and progenitor cells; B, B cells; LEC, lymphatic endothelial cells; NK, natural killer cells; SMC, smooth muscle cells; and T, T cells. (d) A volcano plot depicts differential expression (DE) of the unique adipocyte marker genes by tissue. Genes are grouped into marker genes specific to SAT (MGSS), specific to VAT (MGSV), and shared between the SAT and VAT tissues (MGSBT) (see Methods), and then colored by these groups.

### Marker gene analysis reveals distinct differences between SAT and VAT in cell-type level expression profiles

To elucidate cell-type level functional differences between SAT and VAT, the two main human fat depots^1^, we first investigated SAT and VAT gene expression profiles at the cell-type resolution using SAT and VAT biopsies from the same individuals (Figure 1b-c). These dual tissue fat depot data from the same individuals, which reduce confounding by phenotypic differences between the donors of the SAT and VAT biopsies, enabled us to identify unique SAT and VAT cell-type marker genes, or genes that are differentially upregulated in only one cell-type, and also unique to each adipose tissue depot in their marker gene status (see Methods). Figure 1d shows the adipocyte cell-type markers distributed by tissue specificity. Supplementary Table 1 shows the identified unique cell-type marker genes for all SAT and VAT cell-types after integrating the single nucleus level data of the two tissues. As small differences often drive depot-specific gene and functional patterns^42^, we sought to study not only differences in expression between the depots but also shared transcriptional profiles, as this may elucidate the connection and shared characteristics between the tissues. Thus, from each cell-type marker set, we identified sets of unique marker genes specific to each tissue and shared across the two, resulting in 3 sets: marker genes specific to SAT (MGSS), marker genes specific to VAT (MGSV), and marker genes shared between tissues (MGSBT) (Supplementary Table 2). We focused our analyses on the three major shared cell-types between SAT and VAT, i.e., adipose stem and progenitor cells (ASPCs), adipocytes, and macrophages due to their functional pathway enrichments (see below) and highest number of nuclei available across the two tissues. The MGSS sets contained 244, 208, and 250 unique cell-type marker genes for ASPCs, adipocytes, and macrophages; the MGSV sets 131, 225, and 224; and the MGSBT sets 112, 155, and 9, respectively (Supplementary Table 2).

### Shared and tissue-specific marker genes are enriched for genetic signals of obesity and related CMDs

As both adipose tissue depots are known to be impacted by obesity and related disorders^2,4,5,54^, we first examined these marker gene sets for GWAS variant enrichments of obesity and related CMDs. Accordingly, we tested the variants in the *cis* regions (±500kb) of the three sets per cell-type in three cell-types, MGSS, MGSV, and MGSBT (nine total sets), for enrichment of BMI, abdominal obesity (using WHRadjBMI as a proxy), and type 2 diabetes (T2D) GWAS signals using GWAS summary statistics and the MAGENTA tool (see Methods). We observed significant enrichments (FDR<0.05) in the ASPC MGSBT for BMI, and in the adipocyte MGSS for T2D and WHRadjBMI (Table 1), while no enrichments were detected for any cell-types with the MGSV sets.

**Table 1.** Variants in the *cis* regions of genes in the SAT and tissue-shared adipocyte unique marker gene sets are significantly enriched for obesity and type 2 diabetes GWAS SNPs.

| Gene set<sup>A</sup> | GWAS outcome<sup>B</sup> | FDR<sup>C</sup> |
| --- | --- | --- |
| Adipocyte<br>MGSS | T2D | $4.30 \times 10^{-3}$ |
| Adipocyte<br>MGSS | WHRadjBMI | $1.77 \times 10^{-2}$ |
| ASPC<br>MGSBT | BMI | $2.93 \times 10^{-2}$ |

<sup>A</sup> MGSS indicates the unique marker genes specific to subcutaneous adipose tissue (SAT), and MGSBT the unique marker genes shared between the two fat depots.
<sup>B</sup> WHRadjBMI indicates waist-hip-ratio adjusted for body mass index, T2D type 2 diabetes, BMI body mass index, and GWAS genome-wide association study.
<sup>C</sup> Significance was determined by comparing observed vs expected number of genes that passed 75% enrichment cutoff using MAGENTA<sup>38</sup>.

Next, we assessed each marker gene set for enrichment of polygenic risk for these obesity and related traits. We evaluated the predictive power of annotated polygenic risk scores (PRSs) for WHRadjBMI, BMI, and T2D from the *cis* regional variants of each set in the large UK Biobank (UKB) cohort. We first built PRSs for all individuals for each cell-type set and found that the regional PRS constructed for the adipocyte MGSS gene set was a significant predictor (p_R_^2^<2.23×10^-308^) of WHRadjBMI. Next, we followed up on these WHRadjBMI PRS findings using permutations. Of our 10,000 permutation scores, each similarly built from SNPs residing in the *cis* regions of the same number of randomly selected adipocyte expressed genes from either tissue, we observed only 0.82% to have an incremental R^2^ greater than or equal to that of the adipocyte MGSS WHRadjBMI PRS (R^2^=1.20%; p_perm10,000_=0.0082) (Figure 2; Supplementary Table 3). Due to this observed enrichment in variance explained of WHRadjBMI, which is known to be a sex-dimorphic trait^43,55^, we also constructed these regional WHRadjBMI PRSs for females and males separately. We observed the enrichment in all individuals to be driven by females as the WHRadjBMI PRS constructed for the *cis* regional variants of the adipocyte MGSS genes was significantly enriched (R^2^=2.12%, p_perm10,000_= 0.0101) in females, but not in males (Figure 2; Supplementary Table 3). In line with previous connections with visceral adipose tissue^56,57^, we found that the variance explained for T2D was significantly enriched for the adipocyte MGSV gene set (Delta AUC=1.16%, p_perm1,000_= 0.036). No other significant enrichments in variance explained were detected for BMI or T2D in our PRS analyses, and no other significant enrichments were detected for other cell-types for WHRadjBMI.

**Figure 2.**
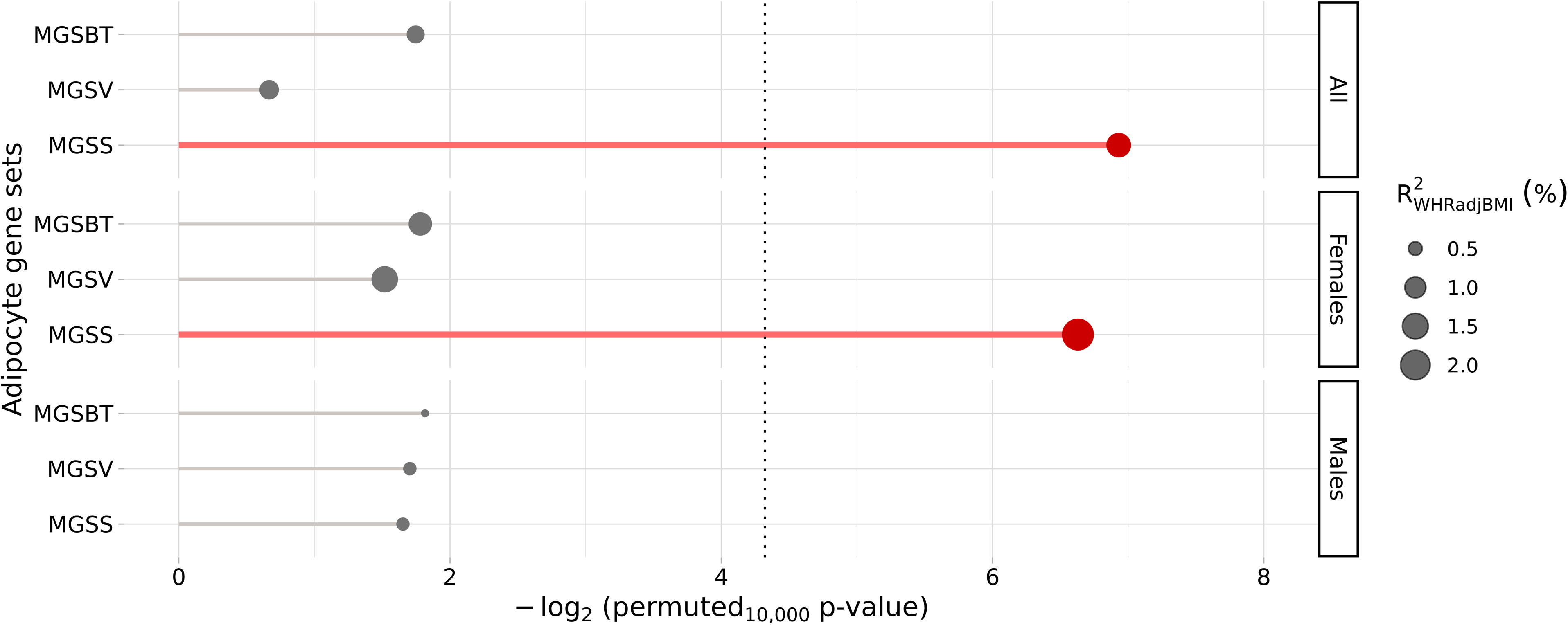
Regional polygenic risk scores (PRSs) constructed from the *cis* regional variants of the adipocyte MGSS gene sets show enrichment in variance explained for abdominal obesity in all individuals and females of the UK Biobank. Lollipop plots visualize the predictive performance of each regional PRS relative to the 10,000 PRSs built using the same number of randomly selected adipocyte-expressed genes. Each horizontal lollipop represents a separate regional PRS, broken into three categories on the left (all individuals, females, and males). Dot size corresponds to the variance explained, R^2^, of each PRS. The vertical dotted line represents the cutoff for significance (p_perm10,000_<0.05).

After identifying the adipocyte MGSS WHRadjBMI PRS results as significantly enriched for variance explained for all individuals and females, we performed a partitioned heritability analysis to further corroborate these enrichments. We observed that the heritability (h^2^) of WHRadjBMI from the SNPs in the *cis* regions of the adipocyte MGSS set was significantly enriched in all individuals (h^2^ enrichment=2.21, p=0.00150) and in females (h^2^ enrichment=2.47, p=1.54× 10^-4^), in line with the regional PRS analysis.

Taken together, the GWAS and PRS enrichments and significant heritability enrichment results highlight the adipocyte MGSS gene set as particularly important for the polygenic risk of abdominal obesity.

### Identification of *SREBF1* as the key SAT-enriched adipocyte marker gene

After identifying WHRadjBMI PRS enrichments in the adipocyte MGSS set, we explored the functional differences of the SAT and VAT gene sets. For each major cell-type, we tested for overrepresentation of genes in Gene Ontology (GO) categories using all three sets, MGSS, MGSV, and MGSBT (Supplementary Tables 4-10). There were no significant functional pathways for the macrophage MGSS and MGSBT sets. Among the adipocyte MGSV and MGSBT gene sets, we detected 170 and 9 significant pathways, respectively (FDR<0.05). Among the pathways of the adipocyte MGSV gene set, 103 (61%) and 98 (58%) pathways included the well-known insulin metabolism genes, *INSR* and *IRS2*, respectively. However, the strongest adipose tissue function-related pathway enrichments were observed in the adipocyte MGSS set (FDR<0.05), including regulation of fatty acid metabolic process, positive regulation of triglyceride metabolic process, and neutral lipid metabolic process amongst other breakdown-of-molecule centered processes (Supplementary Table 4).

In more detail, we found 23 significant functional pathways (FDR<0.05) for the adipocyte MGSS set (Figure 3a; Supplementary Table 4). Notably, we observed a strong presence of the master transcription factor (TF) of adipogenesis and fatty acid biosynthesis, *SREBF1*^14^, in these MGSS pathways. Of the 23 enriched functional pathways for adipocyte MGSSs (FDR< 0.05), 20 (87%) included *SREBF1*, making it the most prevalent pathway gene, and thus supporting its importance in the SAT adipocyte function. Consistent with the known role of *SREBF1* as an adipose master TF^14,58,59^, we also observed that 8 of the most prevalent pathway genes (genes present in 8 or more pathways) in the adipocyte MGSS gene set are predicted transcription targets of *SREBF1* that have also previously been linked to obesity, including *LEP*^60^ and *FASN*^61^ (Figure 3b).

**Figure 3.**
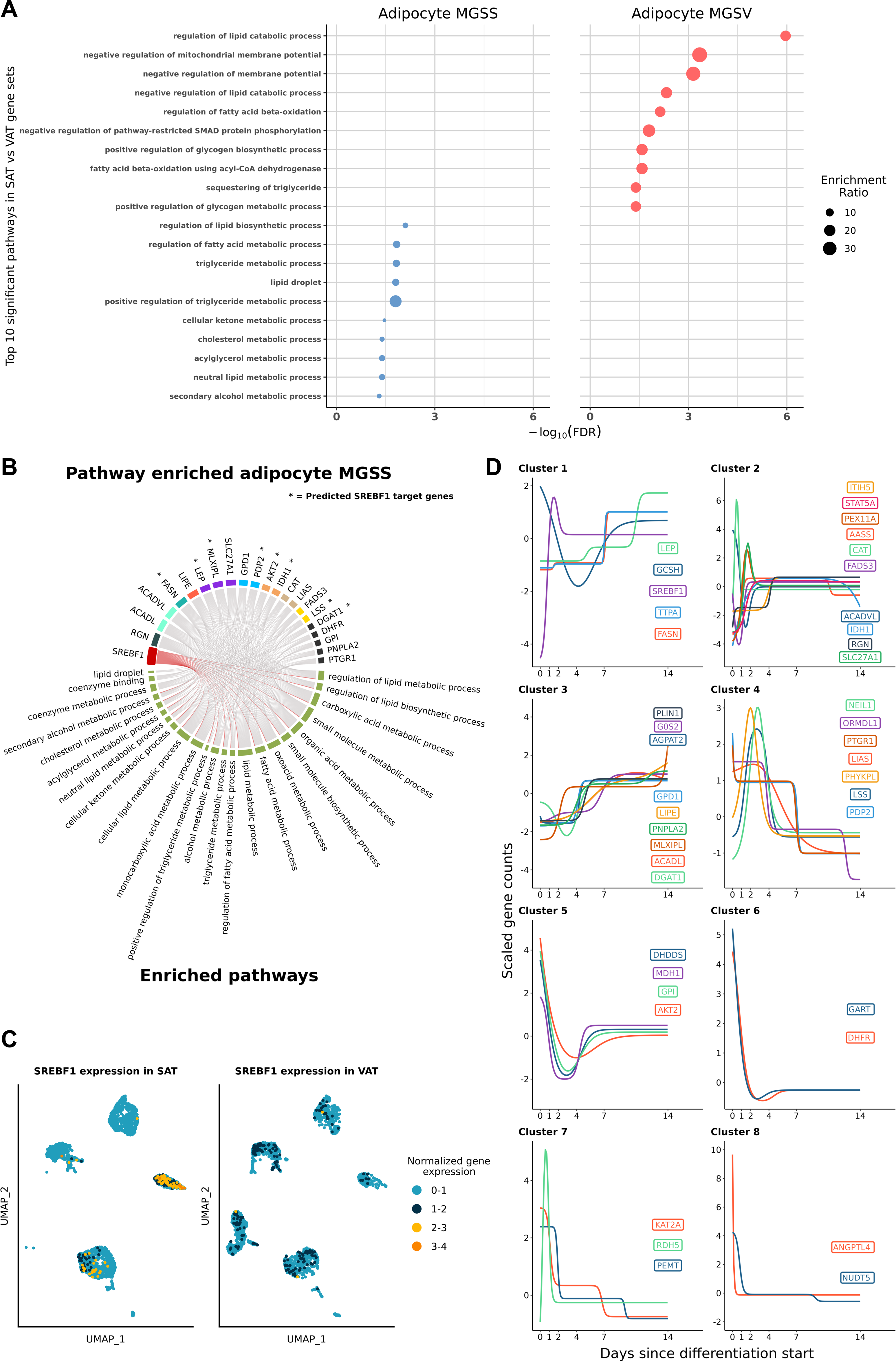
Functional pathway enrichment analyses of cell- and tissue-type level expression profiles reveal distinctive differences in SAT and VAT pathways and highlight the adipose master transcription factor *SREBF1* as the central pathway gene present in 87% of the adipocyte MGSS pathways. (a) The top 10 most significant (FDR<0.05) functional pathways for adipocyte MGSS and MGSV are shown side by side, colored by adipocyte tissue-specific set. P-values for each pathway are shown on the x-axis, and each pathway is subsequently ranked by decreasing p-value. The point size of each dot corresponds to the enrichment ratio of the pathway. (b) Circos plots visualize the pathway memberships of the adipocyte MGSS genes that appear in >=8 adipocyte MGSS pathways and connect the genes to their respective pathways. Genes denoted with an asterisk are predicted target genes of *SREBF1*. Each gene is colored by the number of pathways it belongs to. We highlight *SREBF1*, an adipose tissue master TF and a key pathway gene that is present in 20 out of 23 pathways enriched in adipocyte MGSSs. (c) The cell-type level expression of *SREBF1* across both fat depots is shown on the UMAP space, separately for SAT and VAT, and colored by normalized gene expression counts. d) The scaled expression counts over a 14-day adipocyte differentiation experiment of the longitudinally differentially expressed (DE) adipocyte MGSS genes with memberships in multiple enriched pathways are plotted and grouped by their longitudinal expression trajectory clusters.

Figure 3c shows how this key MGSS TF, *SREBF1* exhibits a substantially higher adipocyte-enriched expression level in SAT than VAT. We confirmed the adipocyte unique marker gene status of *SREBF1* in SAT snRNA-seq in the independent RYSA cohort (n=68) (average log_2_ fold change=0.473, p_adj_<2.23×10^-308^), thus replicating its role as a significant SAT specific marker. Additionally, we replicated the lack of preferential expression of *SREBF1* in VAT adipocytes, in an independent VAT snRNA-seq data from another cohort^34^ comprising obese individuals (BMI>30) of European descent (n=5), which, similarly to the KOBS VAT samples, showed low, diffuse expression of *SREBF1* in all VAT cell-types (Supplementary Figure 2).

### Identification of temporally co-expressed adipocyte MGSS pathway genes during human adipogenesis

To link these functionally enriched MGSS genes to adipose tissue development and differentiation, we examined the temporal expression of the key pathway genes within the adipocyte MGSS set during human primary SAT preadipocyte differentiation (i.e., adipogenesis). We first identified 43 MGSS genes that contributed to more than one significantly (FDR<0.05) overrepresented pathway (Supplementary Table 4) and then found that the 42 of the 43 genes, expressed during SAT adipogenesis, were all longitudinally differentially expressed (DE) (FDR<0.05) during differentiation (Figure 3d). To link together genes with similar temporal transcription patterns, we clustered these longitudinally expressed genes using DPGP^51^ into 8 groups of temporally co-expressed genes (Figure 3d, Supplementary Table 11). Of note, we observed *SREBF1* to cluster with two previously described obesity genes, *LEP*^60^ and *FASN*^61^ (Figure 3d), further supporting the coordinated role of *SREBF1* as a master TF with other key adipocyte marker genes.

### Longitudinally differentially accessible peaks, co-accessible in the *SREBF1* region during adipogenesis, harbor abdominal obesity GWAS variants

To elucidate the genetic regulatory role of *SREBF1* in abdominal obesity, we first searched for open chromatin peaks with similar temporal accessibility patterns in the *cis* region of *SREBF1* during human SAT adipogenesis (Figure 4a). We detected 129 peaks in the *cis* region (± 500 kb) of *SREBF1* and found 120 (93%) to be longitudinally DA over the 14-day preadipocyte differentiation data (FDR<0.05), which we then clustered into 10 groups using DPGP^51^ (Supplementary Table 12). The clusters with peaks remaining after thresholding at a cluster assignment probability>0.9 are shown in Supplementary Figure 3. We focused on cluster 4, which contains 5 peaks with similar trajectories over the 14-day differentiation (Figure 4b).

**Figure 4.**
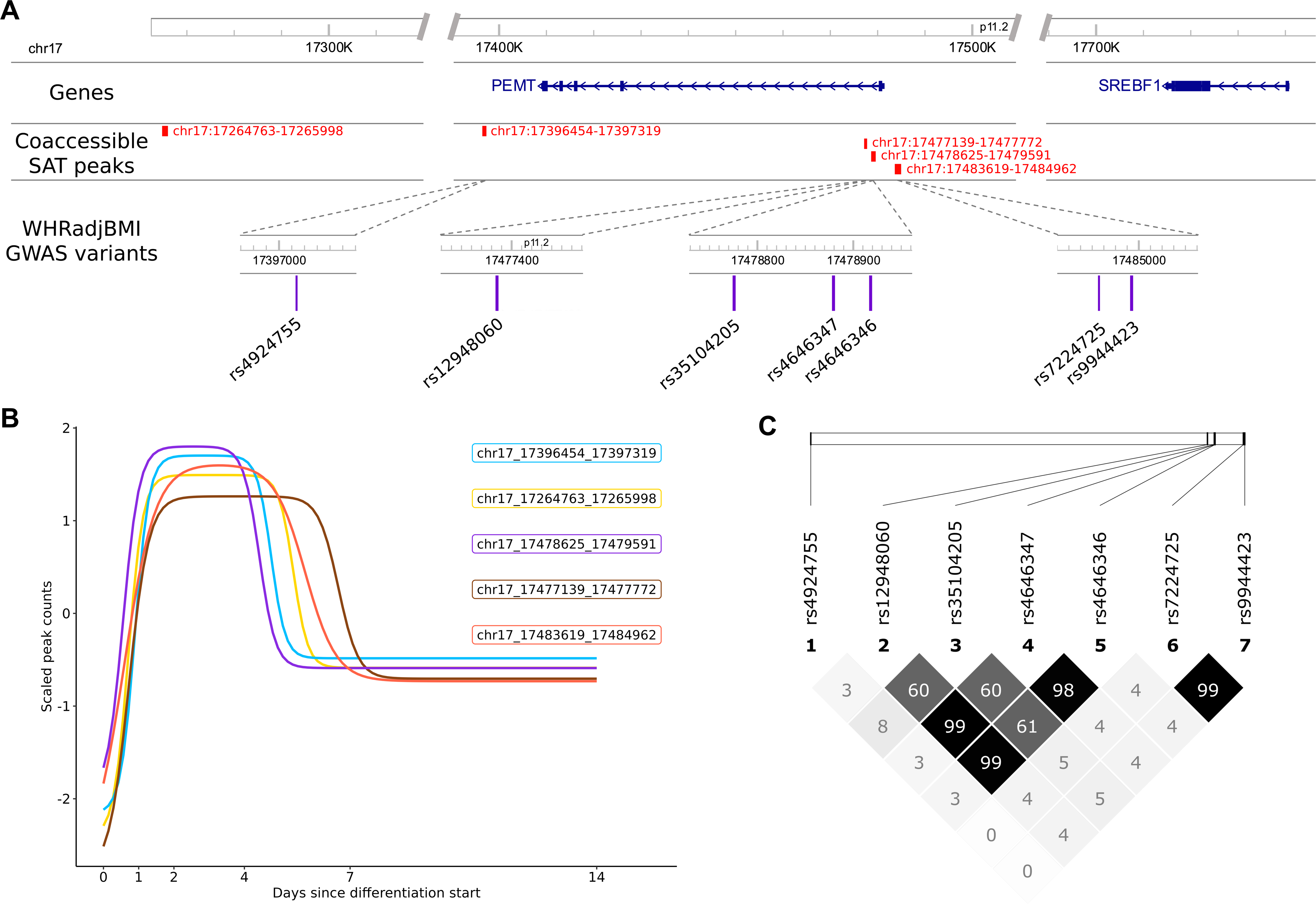
Longitudinal trajectory analysis of open chromatin reveals co-accessible peaks in the *cis* region of *SREBF1* that harbor WHRadjBMI variants. (a) A regional schematic overview shows the proximity to *SREBF1* of 5 co-accessible peaks that exhibit very similar open chromatin trajectory patterns. The peaks collectively harbor WHRadjGWAS variants, viewable through zoomed in loci of the chromosome. (b) The temporal accessibility patterns of the 5 peaks in the *SREBF1* are plotted across the 14-day, 6 time-point SAT preadipocyte differentiation experiment. (c) The pairwise linkage disequilibrium (LD) (R^2^) in the UK Biobank (n=9,981) between the 7 WHRadjBMI GWAS SNPs residing within the co-accessible adipocyte peaks in the *SREBF1 cis* region is shown as a HaploView plot, colored by R^2^.

To determine whether these five co-accessible peaks from cluster 4 in the *SREBF1* region (Figure 4b) are involved in genetic risk of abdominal obesity, we next investigated them for overlaps with WHRadjBMI GWAS variants. Using the large, previously published GWAS summary statistics for WHRadjBMI from GIANT-UKB^52^, we identified 7 GWAS SNPs within these 5 peaks (Table 2). Of the 7 SNPs, rs4924755, rs7224725, and rs9944423 were significant (p<5×10^-8^) in all individuals, driven by females (also significant in only females); rs35104205 was significant in all, females, and males; and rs12948060, rs4646347, and rs4646346 were significant only in males, reflecting the well-known sex effects of the WHRadjBMI trait (Table 2). Furthermore, rs4924755, rs7224725, rs9944423, and rs35104205 are also significant GWAS SNPs (p<5×10^-8^) for the serum total triglycerides in all individuals from the European ancestry GLGC lipid GWAS^62^.

**Table 2.**
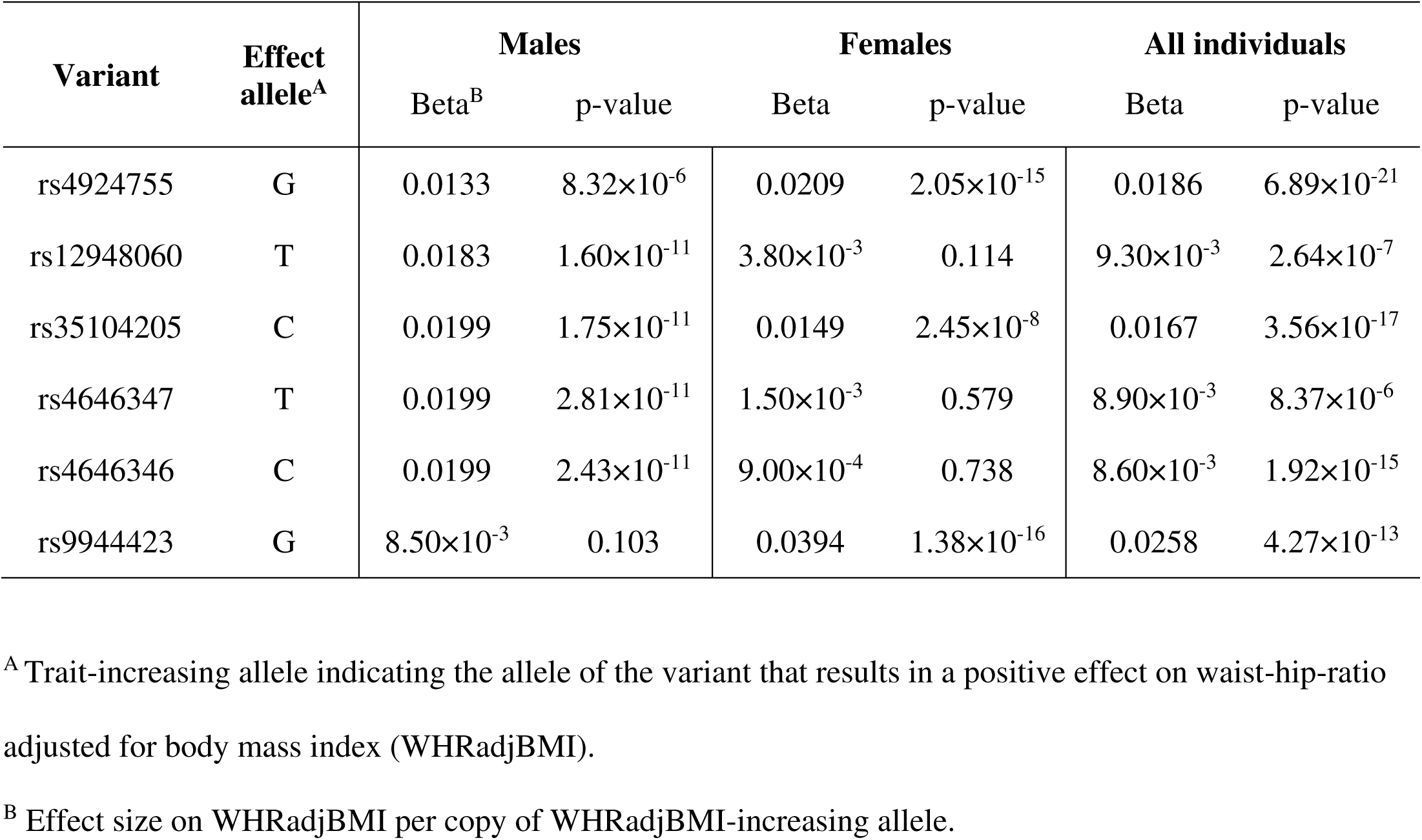
Open chromatin peaks co-accessible during human SAT adipogenesis in the *cis* region of *SREBF1* harbor seven genome-wide significant WHRadjBMI GWAS variants from GIANT-UKB WHRadjBMI GWAS^52^.

| Variant | Effect allele<sup>A</sup> | Males |  | Females |  | All individuals |  |
| --- | --- | --- | --- | --- | --- | --- | --- |
|  |  | Beta<sup>B</sup> | p-value | Beta | p-value | Beta | p-value |
| rs4924755 | G | 0.0133 | 8.32×10<sup>-6</sup> | 0.0209 | 2.05×10<sup>-15</sup> | 0.0186 | 6.89×10<sup>-21</sup> |
| rs12948060 | T | 0.0183 | 1.60×10<sup>-11</sup> | 3.80×10<sup>-3</sup> | 0.114 | 9.30×10<sup>-3</sup> | 2.64×10<sup>-7</sup> |
| rs35104205 | C | 0.0199 | 1.75×10<sup>-11</sup> | 0.0149 | 2.45×10<sup>-8</sup> | 0.0167 | 3.56×10<sup>-17</sup> |
| rs4646347 | T | 0.0199 | 2.81×10<sup>-11</sup> | 1.50×10<sup>-3</sup> | 0.579 | 8.90×10<sup>-3</sup> | 8.37×10<sup>-6</sup> |
| rs4646346 | C | 0.0199 | 2.43×10<sup>-11</sup> | 9.00×10<sup>-4</sup> | 0.738 | 8.60×10<sup>-3</sup> | 1.92×10<sup>-15</sup> |
| rs9944423 | G | 8.50×10<sup>-3</sup> | 0.103 | 0.0394 | 1.38×10<sup>-16</sup> | 0.0258 | 4.27×10<sup>-13</sup> |

<sup>A</sup> Trait-increasing allele indicating the allele of the variant that results in a positive effect on waist-hip-ratio adjusted for body mass index (WHRadjBMI).
<sup>B</sup> Effect size on WHRadjBMI per copy of WHRadjBMI-increasing allele.

**Table 3.**
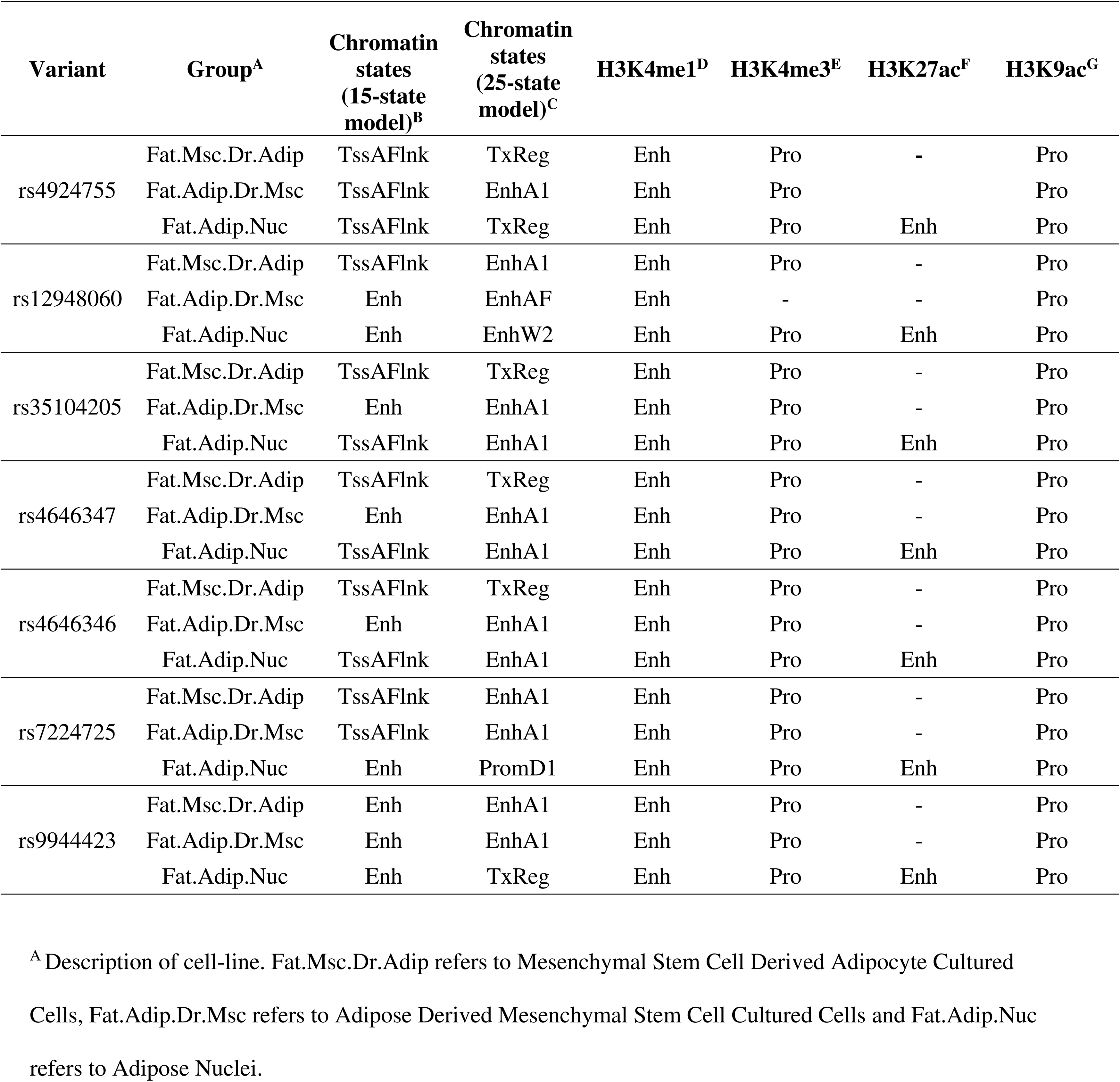
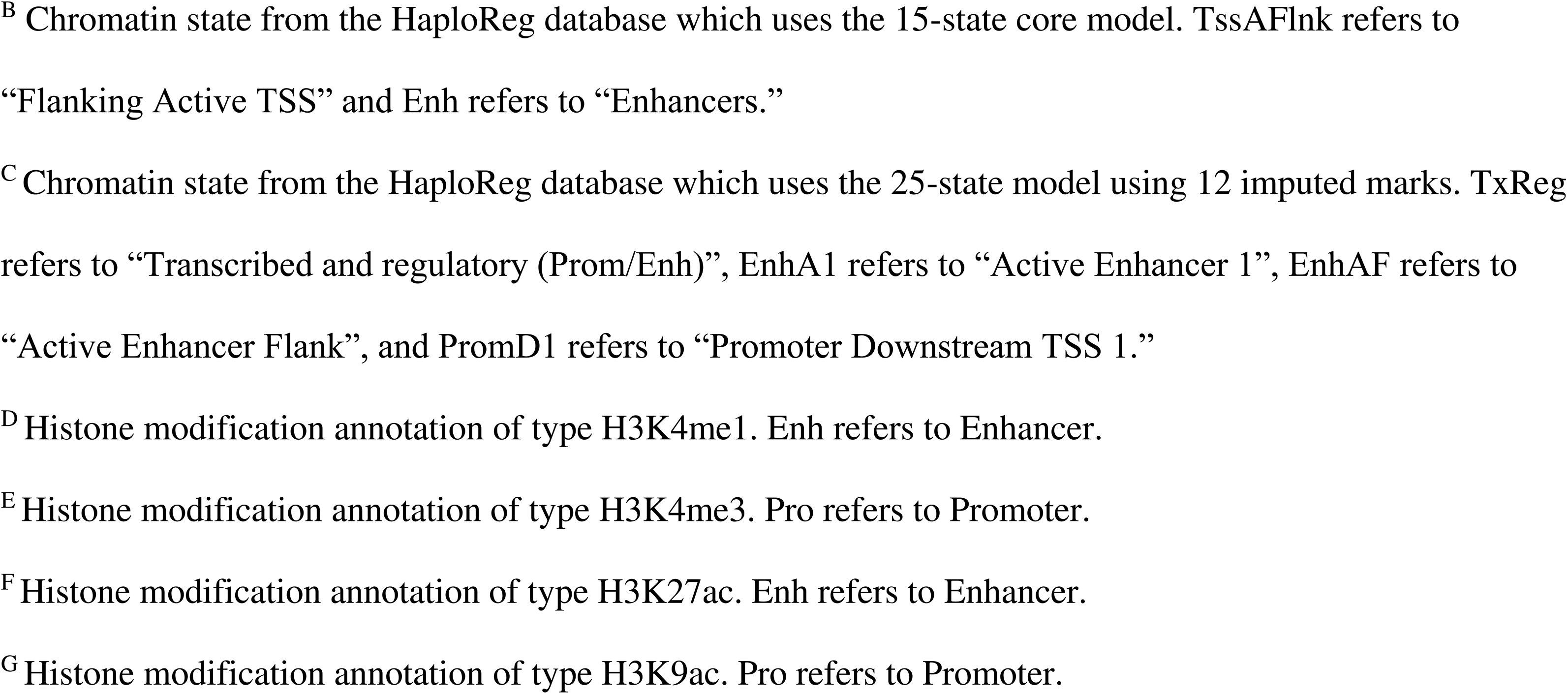
Investigation of seven WHRadjBMI GWAS variants for histone modifications reveals activity in enhancer and promoter regions.

| Variant | Group<sup>A</sup> | Chromatin states (15-state model)<sup>B</sup> | Chromatin states (25-state model)<sup>C</sup> | H3K4me1<sup>D</sup> | H3K4me3<sup>E</sup> | H3K27ac<sup>F</sup> | H3K9ac<sup>G</sup> |
| --- | --- | --- | --- | --- | --- | --- | --- |
| rs4924755 | Fat.Msc.Dr.Adip | TssAFlnk | TxReg | Enh | Pro | - | Pro |
|  | Fat.Adip.Dr.Msc | TssAFlnk | EnhA1 | Enh | Pro |  | Pro |
|  | Fat.Adip.Nuc | TssAFlnk | TxReg | Enh | Pro | Enh | Pro |
| rs12948060 | Fat.Msc.Dr.Adip | TssAFlnk | EnhA1 | Enh | Pro | - | Pro |
|  | Fat.Adip.Dr.Msc | Enh | EnhAF | Enh | - | - | Pro |
|  | Fat.Adip.Nuc | Enh | EnhW2 | Enh | Pro | Enh | Pro |
| rs35104205 | Fat.Msc.Dr.Adip | TssAFlnk | TxReg | Enh | Pro | - | Pro |
|  | Fat.Adip.Dr.Msc | Enh | EnhA1 | Enh | Pro | - | Pro |
|  | Fat.Adip.Nuc | TssAFlnk | EnhA1 | Enh | Pro | Enh | Pro |
| rs4646347 | Fat.Msc.Dr.Adip | TssAFlnk | TxReg | Enh | Pro | - | Pro |
|  | Fat.Adip.Dr.Msc | Enh | EnhA1 | Enh | Pro | - | Pro |
|  | Fat.Adip.Nuc | TssAFlnk | EnhA1 | Enh | Pro | Enh | Pro |
| rs4646346 | Fat.Msc.Dr.Adip | TssAFlnk | TxReg | Enh | Pro | - | Pro |
|  | Fat.Adip.Dr.Msc | Enh | EnhA1 | Enh | Pro | - | Pro |
|  | Fat.Adip.Nuc | TssAFlnk | EnhA1 | Enh | Pro | Enh | Pro |
| rs7224725 | Fat.Msc.Dr.Adip | TssAFlnk | EnhA1 | Enh | Pro | - | Pro |
|  | Fat.Adip.Dr.Msc | TssAFlnk | EnhA1 | Enh | Pro | - | Pro |
|  | Fat.Adip.Nuc | Enh | PromD1 | Enh | Pro | Enh | Pro |
| rs9944423 | Fat.Msc.Dr.Adip | Enh | EnhA1 | Enh | Pro | - | Pro |
|  | Fat.Adip.Dr.Msc | Enh | EnhA1 | Enh | Pro | - | Pro |
|  | Fat.Adip.Nuc | Enh | TxReg | Enh | Pro | Enh | Pro |

<sup>A</sup> Description of cell-line. Fat.Msc.Dr.Adip refers to Mesenchymal Stem Cell Derived Adipocyte Cultured Cells, Fat.Adip.Dr.Msc refers to Adipose Derived Mesenchymal Stem Cell Cultured Cells and Fat.Adip.Nuc refers to Adipose Nuclei.
<sup>B</sup> Chromatin state from the HaploReg database which uses the 15-state core model. TssAFlank refers to “Flanking Active TSS” and Enh refers to “Enhancers.”
<sup>C</sup> Chromatin state from the HaploReg database which uses the 25-state model using 12 imputed marks. TxReg refers to “Transcribed and regulatory (Prom/Enh)”, EnhA1 refers to “Active Enhancer 1”, EnhAF refers to “Active Enhancer Flank”, and PromD1 refers to “Promoter Downstream TSS 1.”
<sup>D</sup> Histone modification annotation of type H3K4me1. Enh refers to Enhancer.
<sup>E</sup> Histone modification annotation of type H3K4me3. Pro refers to Promoter.
<sup>F</sup> Histone modification annotation of type H3K27ac. Enh refers to Enhancer.
<sup>G</sup> Histone modification annotation of type H3K9ac. Pro refers to Promoter.

We observed tight LD patterns (R^2^>99%) among three of the 7 SNPs, rs12948060, rs4646347, and rs4646346, and separately among two SNPs rs7224725 and rs9944423, indicating 4 separate regional WHRadjBMI signals (Figure 4c). These results show that the 5 co-accessible peaks in the *SREBF1* region contribute to the genetic risk of abdominal obesity.

### Risk allele status for the abdominal obesity GWAS SNPs in the *SREBF1 cis* region affects SAT adipocyte expression of *SREBF1* and more than hundred SAT adipocyte marker genes

We hypothesized that these seven abdominal obesity GWAS variants, which reside in the *cis* regulatory region of *SREBF1*, may influence the downstream expression of important adipocyte genes in *trans*. To address this hypothesis, we investigated whether the risk allele status of the seven WHRadjBMI GWAS SNPs in the *SREBF1 cis* region impacts SAT adipocyte expression of *SREBF1* and downstream, the adipocyte expression of other adipocyte MGSSs, using independent SAT snRNA-seq data from another European bariatric surgery cohort, RYSA (n=68) (see Methods). We identified *SREBF1* and 146 other adipocyte MGSS genes, for which the adipocyte expression is significantly affected by the risk allele carrier status of one or more of these seven WHRadjBMI GWAS variants in *trans* (see Methods) (Supplementary Table 13). Due to the tight LD (R^2^>99%) among rs12948060, rs4646347, and rs4646346, as well as among SNPs rs7224725 and rs9944423 (Figure 4c), their results are identical.

In more detail, we observed that the SAT adipocyte expression of *SREBF1* was significantly impacted by each of the seven WHRadjBMI GWAS variants, being higher expressed in the risk allele carriers of rs12948060, rs35104205, rs4646347, and rs4646346, and lower expressed in the risk allele carriers of rs4924755, rs7224725, and rs9944423. The three SNPs in tight LD (rs12948060, rs4646347, and rs4646346) had the largest number of genes (n=115 genes) with significantly higher expression in the risk allele carriers (Supplementary Table 13), while the SNPs rs7224725 and rs9944423 in LD (R^2^>99) had the fewest (n=23 genes) (Supplementary Table 13). We found that the variant rs4924755 impacted the largest number of genes with lower expression in the risk-allele carriers (n=48 genes), including multiple functionally important adipose genes, e.g., *FASN*, *AGPAT2*, *DGAT1*, in line with the downregulation of *SREBF1* in these carriers (Supplementary Table 13). The risk allele carrier status of this variant rs4924755 also associated with upregulation of numerous key adipose genes, such as *PLIN1*, *PLIN4*, and *LIPE* (Supplementary Table 13). We show the functional pathway enrichments of the gene sets affected by the risk allele carrier status in Supplementary Tables 14-16.

We also built module scores in the independent RYSA cohort using the SAT adipocyte expression of the up/down regulated *trans* genes by the risk allele status of the of the *SREBF1* abdominal obesity GWAS SNPs (Figure 5a-b, Supplementary Figure 4). These average expression results of the *trans* genes further demonstrate the significant, wide-spread *trans* effects of the *SREBF1* abdominal obesity GWAS SNPs on SAT adipocyte expression. Taken together, our results suggest that the WHRadjBMI GWAS SNPs in the *cis* region of *SREBF1* affect SAT adipocyte expression of the major adipose tissue TF, *SREBF1*, and downstream of that, the adipocyte expression of 146 genes, comprising 70% of all adipocyte MGSS genes and including multiple key adipocyte MGSS genes.

**Figure 5.**
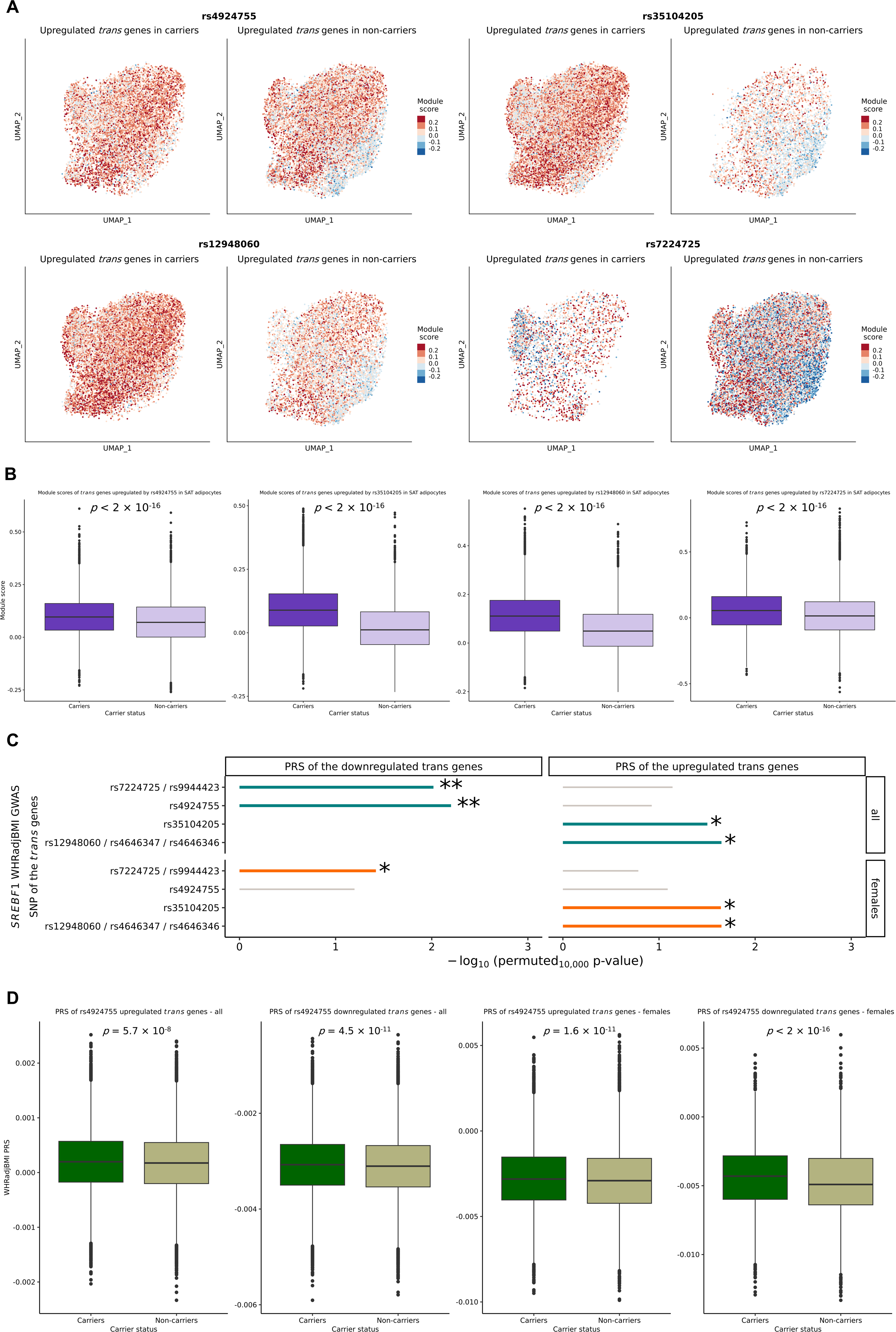
Trans effects of *SREBF1* abdominal obesity GWAS SNPs established using single cell level expression data and biobank investigation. (a) Trans effects of abdominal obesity GWAS SNPs in the *SREBF1* region on SAT adipocyte expression of the adipocyte MGSS genes in an allele-specific way is shown using module scores, constructed in the RYSA cohort (n=68) using the average SAT adipocyte expression of the upregulated *trans* genes by the risk allele status of the *SREBF1* GWAS SNPs. For the module scores of the downregulated *trans* genes, see Supplementary Figure 4. The plots represent the module scores calculated for the upregulated *trans* genes by each SNP (or by a SNP representing a tight LD block, see Figure 4c). Thus, the module scores for the LD block of rs12948060, rs4646347, and rs4646346 and the LD block of rs7224725 and rs9944423 are shown using the data for rs12948060 and rs7224725. Each point represents a cell and its average gene expression of the *trans* genes of interest. (b) The differences between the carriers and non-carriers are also visualized through a two-sided Wilcoxon test and boxplot. (c) The *trans* gene sets are enriched for variance explained in abdominal obesity. Lollipop plots show the regional PRS results for each *tran*s gene set (constructed in all individuals and females). Entries missing a lollipop did not either have enough SNPs to accurately build a PRS (<5 SNPs) or significantly predict the variance in WHRadjBMI (p_R_^2^>0.05) after the regional PRS construction. (c) Differences in the magnitude of the regional PRSs between the carriers and non-carriers of the abdominal obesity risk allele for rs4924755 visualized through boxplots. Significance is calculated using a two-sided Wilcoxon test. Plots for the regional WHRadjBMI PRSs of the *trans* genes calculated in females and all individuals in UKB are displayed.

### Adipocyte open chromatin variants in *cis* regions of the *trans* gene sets are enriched for variance explained in WHRadjBMI and their PRSs differ by the risk allele carrier status of the *SREBF1* abdominal obesity GWAS SNPs

We hypothesized that the *trans* gene sets we identified had downstream effects on the polygenic risk of abdominal obesity by the risk allele carrier status of the *SREBF1* abdominal obesity GWAS SNPs. Accordingly, we first examined the genetic risk contributions from the *trans* genes sets by constructing their regional PRSs for WHRadjBMI in UKB from the SNPs that fell in adipocyte open chromatin peaks in the *cis* regions of each *trans* gene set (see Methods). We found, through 10,000 permutations, that 12 out of the 21 PRSs which were built from variants from upregulated *trans* gene sets were significantly enriched (p_perm10,000_<0.05) for incremental variance explained in abdominal obesity (Figure 5c, Supplementary Table 17), as were 6 out of the 10 PRSs built from variants from downregulated *trans* gene sets (Figure 5c, Supplementary Table 18), indicating that the regulatory regions of these *trans* regulated genes are important for abdominal obesity.

Next, we classified the UKB individuals as the carriers and non-carriers of the seven *SREBF1* abdominal obesity GWAS risk alleles in the same way as in the RYSA adipocyte expression analysis and then compared the WHRadjBMI PRSs of the *trans* gene sets between the carriers and non-carriers in UKB to further assess the *trans* regulatory nature of the carrier status. We found that the *SREBF1 cis* regional variants exhibit significant differences in the magnitude of the regional WHRadjBMI PRSs of 10 out of the 21 upregulated *trans* gene sets between the carriers and non-carriers of the corresponding *SREBF1* abdominal obesity GWAS SNP risk allele (Wilcoxon padj<0.05) (Supplementary Table 17), thus independently confirming the observed *trans* effects in RYSA adipocytes at the population level in UKB. Similarly, we found that 4 out of 10 downregulated *trans* gene sets also show significant PRS differences (Wilcoxon padj< <0.05) between the carriers and non-carriers (Supplementary Table 18). Figure 5d shows these significant differences between the regional WHRadjBMI PRSs of the *trans* genes by the risk allele carrier status of one of the 7 *SREBF1* WHRadjBMI GWAS SNPs, rs4924755, and Supplementary Figure 5 illustrates similar significant WHRadjBMI PRS differences with several of the other SNPs. We also observed sex-specific differences in these PRS results (Figure 5c-d, Supplementary Tables 17-18), in line with the well-known sex differences in abdominal obesity. Overall, our results discover *SREBF1* for SAT adipocyte function and genetic risk of abdominal obesity via variant-specific *trans* effects on numerous adipocyte *trans* genes.

## Discussion

Despite prevailing cellular heterogeneity within and between the two major human fat depots, SAT and VAT, the underlying genes, functional pathways, and epigenetic and genetic factors explaining these differences have remained largely elusive at the cell-type resolution^63–65^. Previous studies have reliably connected SAT and VAT to obesity and related cardiometabolic metabolic diseases (CMDs)^2,4,5,54^, but the distinct contributions of each fat tissue and their cell-types to disease predisposition are less well understood. Here, we integrated human single-cell level RNA data to comprehensively compare SAT and VAT at the transcriptomic level. By deriving tissue- and cell-type-specific marker gene sets (MGSS, MGSV, and MGSBT) for each of the three major adipose cell-types, we establish which unique cell-type marker genes are shared and which ones are specific to adipocytes, ASPCs, and macrophages in SAT versus VAT and how these genes contribute to the key cell-type functions and genetic risks to CMDs. Upon assessing the GWAS associations, partitioned polygenic risk scores, and heritability estimates in these cell-type marker gene sets, we highlighted the importance of the adipocyte marker genes unique to the SAT depot for genetic predisposition to abdominal obesity, led by the major adipose TF gene, *SREBF1*, observed in 87% of the functional pathways unique to the SAT adipocyte marker genes. We also discover regional, longitudinally co-accessible peaks across SAT adipogenesis at this *SREBF1* locus, which harbor seven GWAS variants associated with WHRadjBMI, a well-established abdominal obesity proxy^11^. Next, we show that the risk allele carrier status of these seven WHRadjBMI GWAS variants affects SAT adipocyte expression of *SREBF1* and 146 other functionally enriched SAT adipocyte marker genes (i.e., 70% of all SAT adipocyte marker genes) in the large independent SAT snRNA-seq data set from the RYSA cohort, identifying profound cell-type level downstream effects of this adipose major TF in *trans*. Lastly, we confirm this result independently in the UK Biobank by demonstrating that the partitioned abdominal obesity PRSs of the adipocyte *trans* gene sets differ by the risk allele carrier status of the *SREBF1* abdominal obesity GWAS variants. Taken together, our study discovers abdominal obesity GWAS variants in the *cis* region of *SREBF1* that act in *trans* as drivers of downstream adipocyte gene expression of more than a hundred of genes.

Previous studies have shown that excess VAT leads to increased risk for MASLD, insulin resistance, and coronary artery disease^56,57^ and that excess SAT is correlated with an increase in oxidative stress and inflammation^66^, but the cell-type and gene level mechanisms underlying these different depot-specific contributions to CMD predispositions are not well understood. In our study, by leveraging the snRNA-seq data from the same KOBS participants’ SAT and VAT biopsies, we characterize gene expression without inter-individual bias and separate cell-type marker genes into shared and tissue-specific gene sets. While previous studies have similarly discerned both partial differences in the cell-type identities and functionalities of shared cell-types^67–70^, our study further examines these tissue-shared and specific profiles in the context of the CMD risks. Through constructing partitioned polygenic risk scores which isolate the polygenic risk contributions from the regional variants in the genes at the center of these tissue profiles, we discern enrichment for the abdominal obesity PRS from the adipocyte SAT-specific marker genes, further supported by our WHRadjBMI heritability analysis of the adipocyte MGSS gene set by LD score regression. We additionally confirm previously known connections of VAT to T2D^56,57^ through identifying enrichment for T2D PRS from the adipocyte VAT-specific marker genes. Overall, our genetic results suggest that the genetically regulated transcriptional differences between the two human fat depots link to substantial distinctions between the tissue- and cell-type level contributions to CMDs. When investigating the role of sex as a biological factors, we also found that the regional PRSs constructed from the local variants of the adipocyte SAT-specific marker genes are highly enriched predictors of WHRadjBMI in all individuals and females, in line with the previously established heritability enrichments of WHRadjBMI in SAT, sex-dimorphisms of WHRadjBMI, and the causality estimates of WHRadjBMI for MASLD^71,72^.

We further elucidate patterns of tissue and cell-type specificity among the two fat depots through evaluating the functional enrichments among the SAT and VAT marker gene sets. In line with previously identified cellular heterogeneity between the two tissues^34,70^, we observe that the adipocyte SAT-specific marker genes are enriched for genes involved in the metabolism of fatty acids, triglycerides, and glucose, while the adipocyte marker genes shared across the depots, or specific to VAT exhibited enrichment in molecular movement and lipid localization. These results are supported by previous studies that found that VAT is important for a flux of fatty acid buildup^73^, which suggests that while VAT would be important for the long-term fat storage-oriented properties of adipose tissue, SAT may be more influential for the metabolic activities, such as rapid-expansion of fat molecules, and flexible breakdown of fat-related molecules. In particular, we identified *SREBFI* as the key to the adipocyte MGSS functional pathways, found in 87% of the SAT-specific significant functional pathways. *SREBF1* is the fatty acid synthesis master TF and a key regulator of transcriptional control for adipogenesis^14,15^. It also regulates the transcription of several obesity-involved gene pathways^15^. In our study, we observed a distinct upregulation of *SREBF1* adipocyte expression in SAT, compared to its sparse and diffuse expression across all cell-types in VAT, a pattern that we replicated in independent SAT and VAT cohorts^34^. Consistent with our genetic assessments, these patterns suggest that the contributions to abdominal obesity may be more pronounced in SAT adipocytes than in the VAT counterparts. Although other studies have shown low diffuse expression levels of *SREBF1* across multiple cell-types in VAT^58^, our study discovers *SREBF1* as a unique adipocyte-specific marker gene in SAT involved in numerous functional SAT pathways not enriched among the VAT adipocyte marker genes, which may explain functional differences between SAT and VAT given the well-established regulatory role of this master TF. Furthermore, we present differences in SAT adipocyte gene expression of the key adipocyte marker genes between the carriers and non-carriers of the abdominal obesity GWAS variants in the *cis* region of *SREBF1*, supporting the evidence of *SREBF1* as a master transcription factor with more than a hundred important target genes in SAT adipocytes. Taken together, we find that differences in cell-type-specific gene expression between SAT and VAT link to differences in SAT and VAT function and identify one such gene, *SREBF1* that is present in most SAT adipocyte-specific pathways and absent in VAT adipocyte-specific pathways.

When investigating the genetic contribution of the *SREBF1* region to abdominal obesity, we found that co-accessible adipocyte ATAC-seq peaks in the *cis* region of *SREBF1* harbor multiple genome-wide significant WHRadjBMI GWAS variants, including variants with sex-specific associations, thus linking to the well-established sex-dimorphism of the abdominal obesity^11,43,55^. We also discovered using SAT adipocyte expression data from the large independent RYSA cohort that these WHRadjBMI GWAS risk variants are associated with expression of 146 adipocyte marker genes specific to SAT, including *SREBF1*. These adipocyte marker genes, impacted by the risk allele carrier status, are also functionally enriched for important adipocyte functions. Next, we further confirm these single cell-level *trans* effects by building partitioned abdominal obesity PRSs of these *trans* gene sets in the UK Biobank data, which shows that their partitioned abdominal obesity PRSs differ by the risk allele carrier status of the *SREBF1* abdominal obesity GWAS variants, thus highlighting their downstream *trans* effects of the SREBF1 variants also at the genomic level. Overall, our results identify *trans* effects on SAT adipocyte gene expression by the WHRadjBMI risk variants in the *SREBF1* region using several different data modalities and cohorts. Given the known important TF functional of *SREBF1*, our results support the notion that these WHRadjBMI GWAS risk variants in the regulatory region of *SREBF1* have downstream effects in *trans* on transcriptional regulation of numerous central obesity -related genes. More broadly, our study also provides an integrative genomics approach, leveraging single cell omics and biobank data, that can be generalized to other TFs to accelerate the currently slow discovery of *trans* effects of TF GWAS variants.

We acknowledge several limitations in our study. First, while our KOBS snRNA-seq data pass a rigorous QC, the total number of SAT and VAT nuclei in the KOBS cohort are relatively small. However, the publicly available VAT snRNA-seq data are currently very sparse, and using our dual tissue study design with both SAT and VAT biopsies from the same Finnish individuals with obesity, we at least partially circumvent a well known factor in snRNA-seq data analysis, i.e., the interindividual cellular heterogeneity^34,49^ that may affect the assessment of the cell-type marker genes. Furthermore, we conducted the risk allele carrier status –based analyses in the large independent RYSA snRNA-seq data set and confirmed the key SAT and VAT results of the *SREBF1* cell-type level expression in the RYSA cohort and previous adipose ATLAS by Emont and coworkers^34^. Second, as we assessed adipogenesis data only from the SAT depot due to the well-known practical and technical challenges to obtain viable human VAT primary preadipocytes - which is also reflected by the current lack of longitudinal human VAT adipogenesis RNA-seq data in the public data repositories - it would be important to also explore adipogenesis differentiating human VAT primary preadipocytes in future studies. Third, the cohorts and analyses of this study comprise Europeans from Finland and the UK. Extrapolations to diverse ethnicities are warranted to further investigate these findings. Lastly, the SAT and VAT biopsies in the current study are from individuals with obesity and thus, studies in individuals with normal weight and overweight may provide important additional information regarding the impact of the obese condition on these results.

## Abbreviations

ASPC: Adipose stem and progenitor cell
ATAC: Assay for transposase accessible chromatin
AUC: Area under the curve
BMI: Body mass index
CCA: Canonical correlation analysis
CMD: Cardiometabolic disease
DE: Differential expression
DA: Differential accessibility
GWAS: Genome-wide association study
KOBS: Kuopio Obesity Surgery Study
LD: Linkage disequilibrium
LEC: Lymphatic endothelial cells
MASLD: Metabolic dysfunction-associated steatotic liver disease
MGSBT: Marker genes shared between the SAT and VAT tissues
MGSS: Marker genes specific to SAT
MGSV: Marker genes specific to VAT
NK: Natural killer cells
PCA: Principal component analysis
PRS: Polygenic risk score
RNA-seq: RNA-sequencing
RYSA: Roux-en-Y Gastric Bypass versus Single-Anastomosis Gastric Bypass
SAT: Subcutaneous adipose tissue
SMC: Smooth muscle cells
snRNA-seq: Single nucleus RNA-sequencing
*SREBF1*: Sterol regulatory element-binding transcription factor 1
T2D: Type 2 diabetes
TF: Transcription factor
UKB: UK Biobank
VAT: Visceral adipose tissue
WHRadjBMI: Waist-hip ratio adjusted for BMI

## Author contributions

M. G. S. and P. P. designed the study. M. G. S., A. K., S. H. T. L., K. Z. G., and S. S. D. performed computational analysis and relevant literature comparisons. M. G. S., A. K., S. H. T. L., M. A., and P. P. developed the analytical and statistical approaches. K. M. G. and P. P. generated the ATAC-seq data. U. T. A., S. H. T. L., S. R., K. H. P., M. U. K., and P. P. generated the RNA-seq data. M. A., D. K., and P. P. generated the genotype data. D. K., V. M., H. P., S. H., U. S., T. S., A. J., K. H. P., J. P., and M. U. K. collected the cohort materials. M. G. S., A. K., S. H. T. L., and P. P. wrote the manuscript, and all authors read, reviewed, and/or edited the manuscript.

## Acknowledgments

We would like to thank the participants of the KOBS, RYSA, and Tilkka cohorts, and the UK Biobank. This study was supported by NIH grants R01HL170604 (PP) and R01DK132775 (PP), the Academy of Finland (272376, 266286, 314383, 335443, KHP; 314457 AJ: 338417 SH; and 333021, 335973, MUK), Finnish Medical Foundation (KHP, AJ), Finnish Diabetes Research Foundation (SH, KHP), Orion Foundation (SH), Novo Nordisk Foundation (NNF10OC1013354, NNF17OC0027232, NNF20OC0060547, KHP), Paulo Foundation (SH, KHP), Gyllenberg Foundation (KHP), Sigrid Juselius Foundation (KHP, MUK), Paavo Nurmi Foundation (SH), Helsinki University Hospital Research Funds (SH, KHP, AJ) and the University of Helsinki (KHP). This research was partly supported by the European Research Council (ERC) under the European Union’s Horizon 2020 research and innovation program (Grant 802825 to MUK), and the Finnish Foundation for Cardiovascular Research (MUK). We would like to acknowledge Single Cell Genomics Core and Biocenter Finland for infrastructure support. This work uses data provided by patients and collected by the NHS as part of their care and support. This research was conducted using the UK Biobank Resource under application number 33934.

## Supplemental information

Document S1. Supplementary Table 17, Supplementary Table 18, Supplementary Figure 2, Supplementary Figure 3, Supplementary Figure 4, Supplementary Figure 5

Supplementary Table 1. Excel file containing additional data, related to Figure 1

Supplementary Table 2. Excel file containing additional data, related to Figure 1

Supplementary Table 3. Excel file containing additional data, related to Figure 2

Supplementary Table 4. Excel file containing additional data, related to Figure 3

Supplementary Table 5. Excel file containing additional data, related to Figure 3

Supplementary Table 6. Excel file containing additional data, related to Figure 3

Supplementary Table 7. Excel file containing additional data, related to Figure 3

Supplementary Table 8. Excel file containing additional data, related to Figure 3

Supplementary Table 9. Excel file containing additional data, related to Figure 3

Supplementary Table 10. Excel file containing additional data, related to Figure 3

Supplementary Table 12. Excel file containing additional data, related to Figure 4

Supplementary Table 13. Excel file containing additional data, related to Figure 5

Supplementary Table 14. Excel file containing additional data, related to Figure 5

Supplementary Table 15. Excel file containing additional data, related to Figure 5

Supplementary Table 16. Excel file containing additional data, related to Figure 5

## Data availability statement

The data that support the findings in this manuscript are available from the UK Biobank. However, restrictions apply to the availability of these data, which were used in this study under UK Biobank Application number 33934. UK Biobank data are available for bona fide researchers through the application process: https://www.ukbiobank.ac.uk/learn-more-about-uk-biobank/contact-us. The KOBS VAT snRNA-seq will be made available in the NIH Gene Expression Omnibus (GEO) upon acceptance, under accession number GSEXX. The KOBS SAT snRNA-seq data were made previously available in GEO, under accession number GSE269926. The RYSA SAT snRNA-seq data will be made available in GEO under accession number GSE274778. The data from the primary human preadipocyte differentiation experiment were previously made available in GEO, under accession number GSE249195 for the RNA and GSE269926 for the ATAC. The SAT and VAT snRNA-seq data described in Emont et al.^34^ can be found in the Single Cell Portal under study number SCP1376.

## Code availability

No custom code was used, and all codes used for analyses in this study were unaltered from their publicly available sources, as outlined in the Methods.

