## Supplementary figures and images for "Integration of single cell omics with biobank data discovers *trans* effects of *SREBF1* abdominal obesity risk variants on adipocyte expression of more than 100 genes"

### Supplementary Figure 1

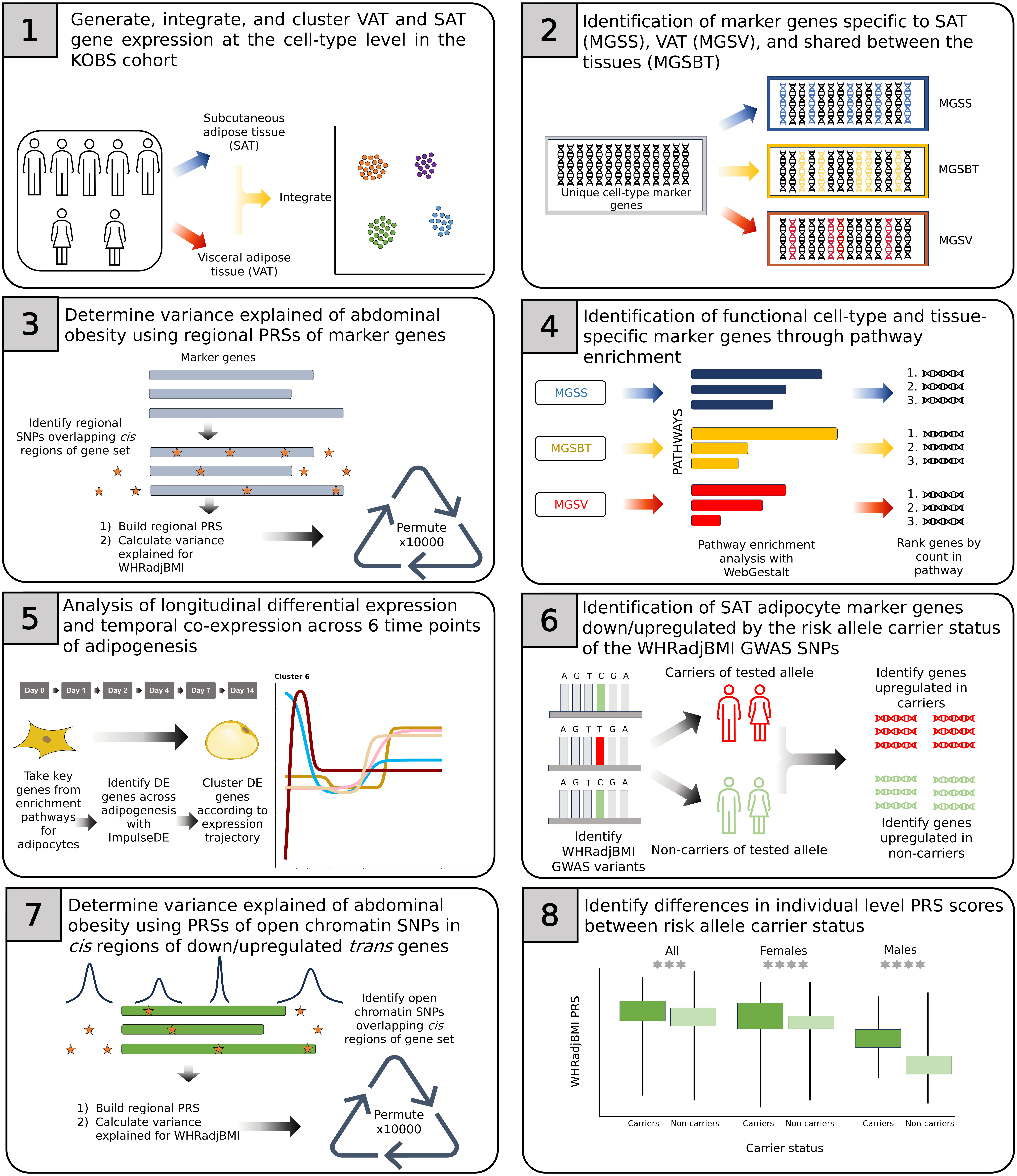
