## Supplementary material for "Integration of single cell omics with biobank data discovers *trans* effects of *SREBF1* abdominal obesity risk variants on adipocyte expression of more than 100 genes": Document S1

**Supplementary figures and tables**

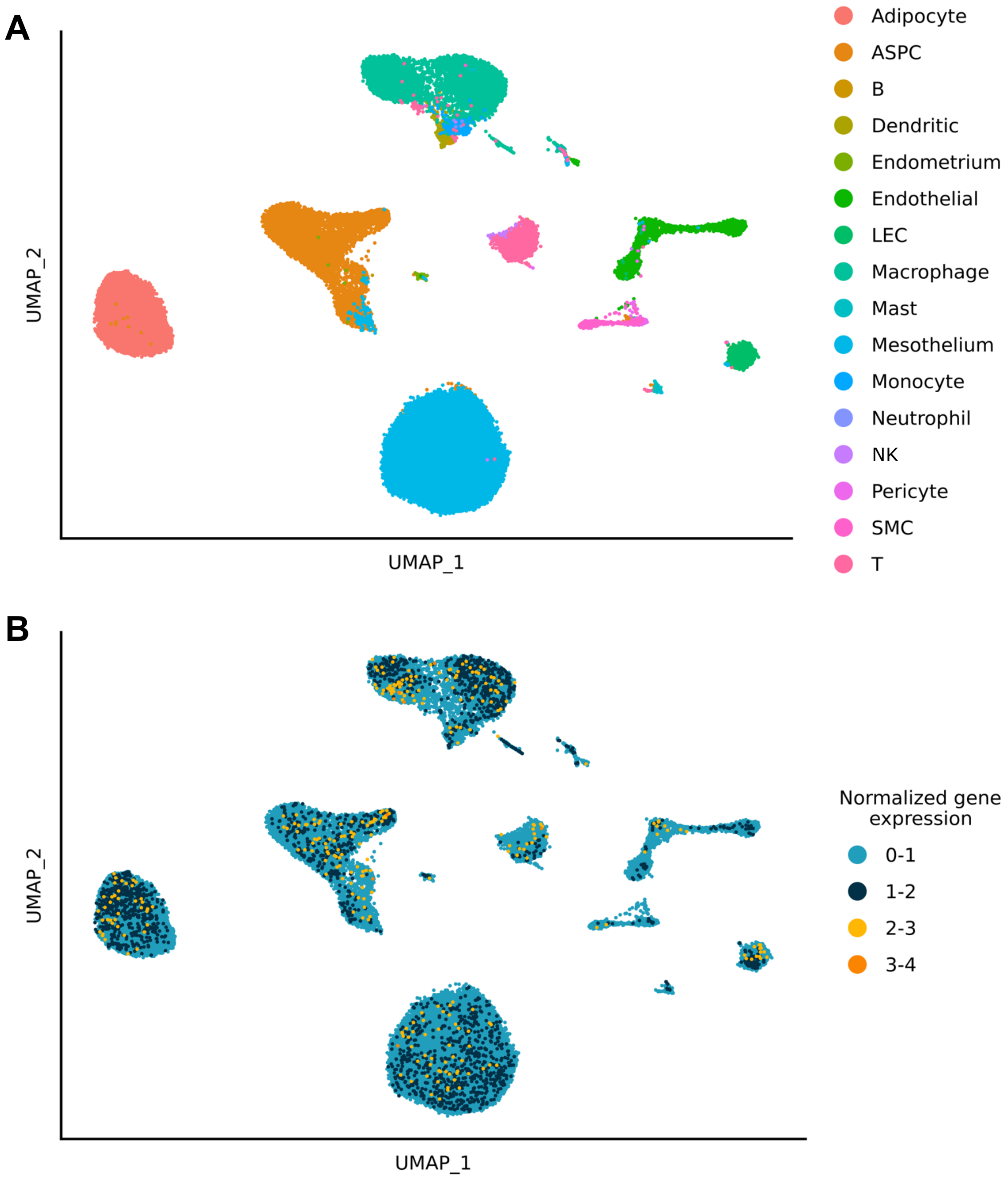


**Supplementary Figure 2. Low, diffuse *SREBF1* gene expression is seen across all cell-types in VAT (n=5 individuals with obesity) using VAT snRNA-seq data from Emont et al.[34]** a) The cluster cell-type identities are visualized on the UMAP space, while mapping each cluster to one of the 16 identified VAT cell-types. ASPC indicates adipose stem and progenitor cells; LEC, lymphatic endothelial cells; NK, natural killer cells; and SMC, smooth muscle cells. b) The VAT cell-type level expression of *SREBF1* is shown on the UMAP space. Each dot represents one cell and each cell is colored by the normalized gene expression counts of *SREBF1*.

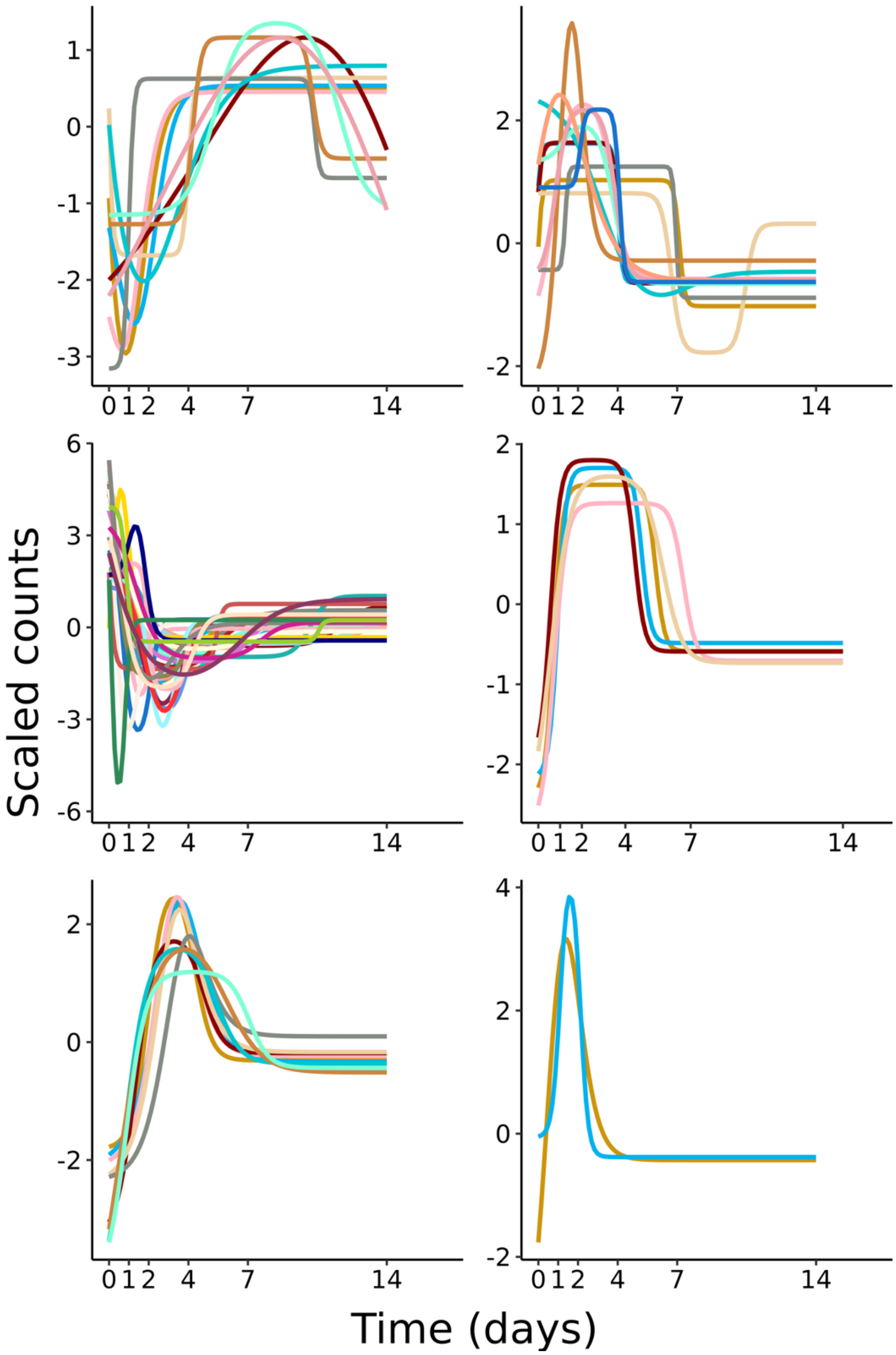


**Supplementary Figure 3. Longitudinal differentially accessible (DA) peaks in the *cis* region of *SREBF1* clustered by temporal accessibility trajectory over human SAT adipogenesis.**

We first identified 129 total peaks in the *cis* region of *SREBF1,* 120 of which are longitudinally DA in temporal ATAC-seq data generated at 6 time-points during the human SAT preadipocyte differentiation. We then assessed the identified peaks for longitudinal co-accessible clusters that have a cluster assignment probability >0.9 using the DPGP tool[51]. These clusters are plotted here. All peaks in this region are listed with their cluster and assignment probabilities in Supplementary Table 12.

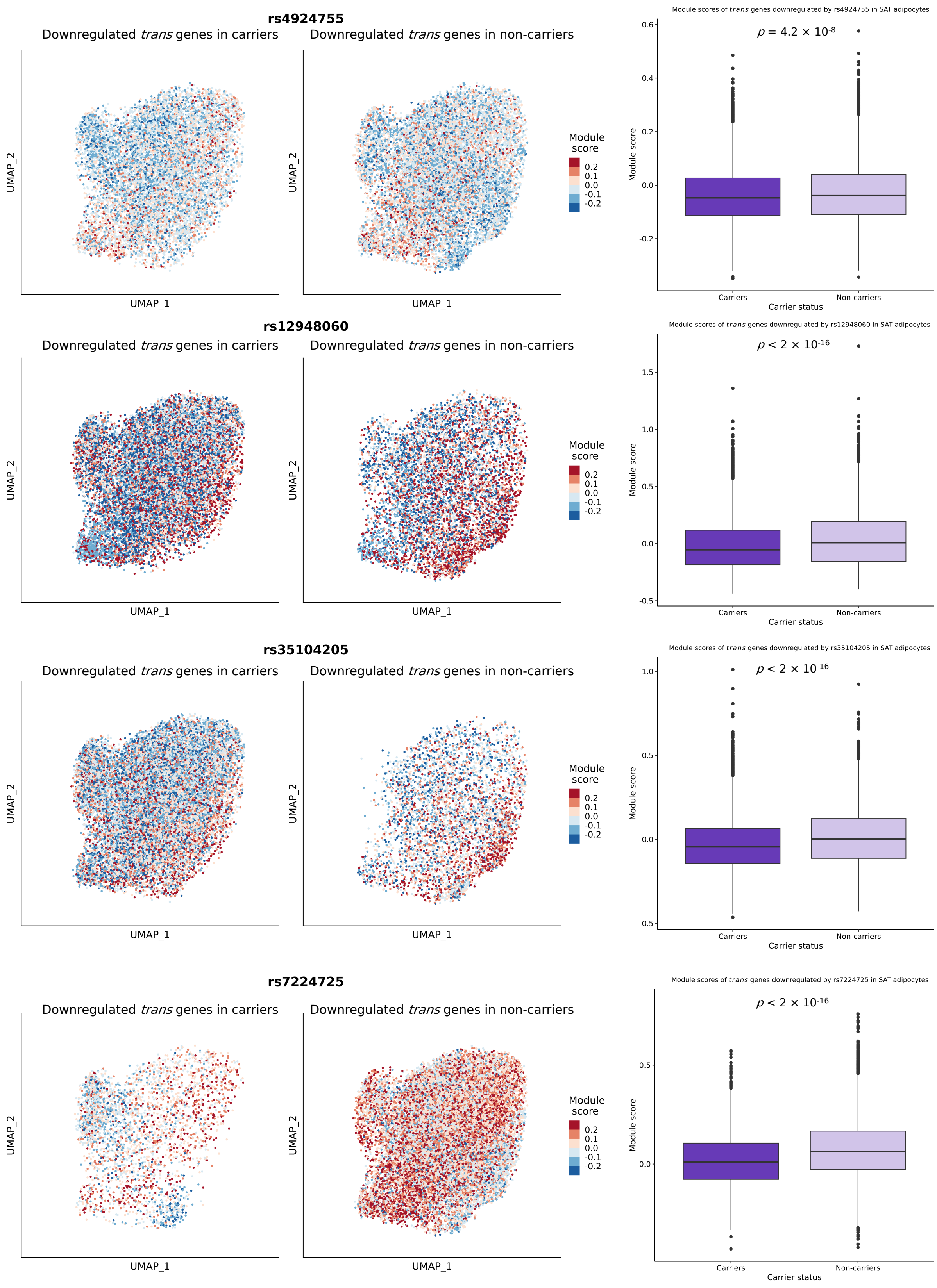

rs4924755
Downregulated trans genes in carriers
Downregulated trans genes in non-carriers
UMAP_1
UMAP_2
Module score
0.2
0.1
0.0
-0.1
-0.2
Module scores of trans genes downregulated by rs4924755 in SAT adipocytes
p = 4.2 × 10-8
Module score
Carriers
Non-carriers
Carrier status
rs12948060
Downregulated trans genes in carriers
Downregulated trans genes in non-carriers
Module scores of trans genes downregulated by rs12948060 in SAT adipocytes
p < 2 × 10-16
rs35104205
Downregulated trans genes in carriers
Downregulated trans genes in non-carriers
Module scores of trans genes downregulated by rs35104205 in SAT adipocytes
p < 2 × 10-16
rs7224725
Downregulated trans genes in carriers
Downregulated trans genes in non-carriers
Module scores of trans genes downregulated by rs7224725 in SAT adipocytes
p < 2 × 10-16

**Supplementary Figure 4.** Module scores in adipocytes of the RYSA cohort using the up/down regulated *trans* gene sets by the risk allele status of each of the 7 SNPs. These plots represent the module scores not shown in Figure 5a and 5b, calculated for either the up- or downregulated *trans* gene set for a SNP or one SNP from each tight LD block (for pair-wise LD between the 7 *SREBF1* WHRadjBMI GWAS SNPs, see Figure 4c). The module scores for the LD block of rs12948060, rs4646347, and rs4646346 and the LD block of rs7224725 and rs9944423 are shown using the data for rs12948060 and rs7224725. Each point represents a cell and its average adipocyte expression of the *trans* genes of interest.

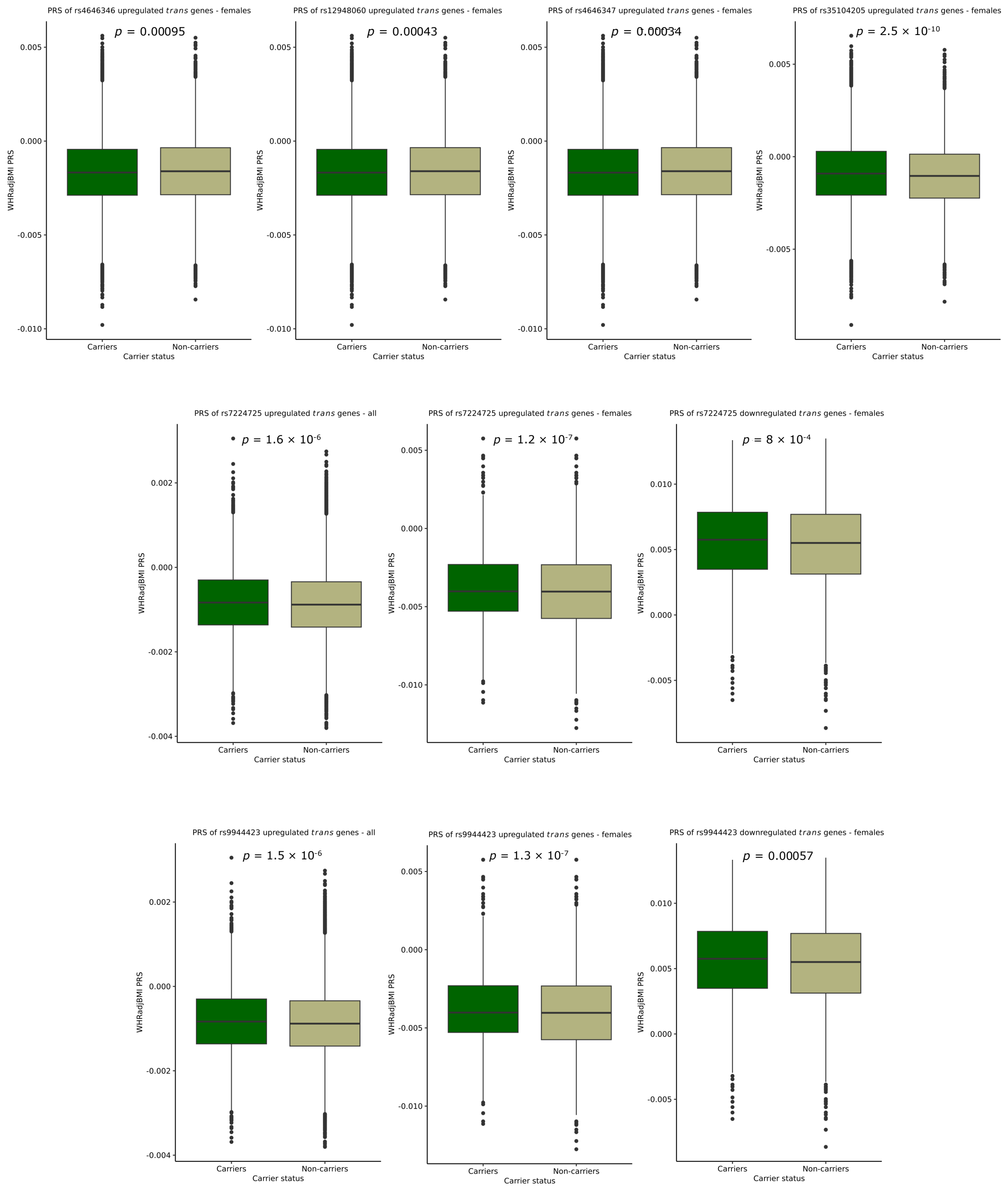


**Supplementary Figure 5.** Significant differences (Wilcoxon-p<0.05) in the magnitude of the regional WHRadjBMI PRSs between the carriers and non-carriers of the risk allele for the remaining *SREBF1* WHRadjBMI GWAS SNPs not shown in Figure 5d, visualized through boxplots.

**Supplementary Table 17.** Variants residing in the adipocyte open chromatin in the *cis* regions of the upregulated *trans* gene sets reveal significantly enriched ($p_{perm}$<0.05) polygenic risk scores (PRSs) for WHRadjBMI and significant differences in the magnitude of their regional WHRadjBMI PRSs between carriers and non-carriers of the trait-increasing risk allele of the seven *SREBF1* WHRadjBMI GWAS SNPs.

| Variant | Effect allele[A] | Group | $R^2_{WHRadjBMI}$ (%)[B] | Permuted p-value[C] | Wilcoxon p-value[D] | Pathway enriched[E] |
|---|---|---|---|---|---|---|
| rs4924755 | G | All | 0.029 | 0.119 (1,000) | $5.66\times10^{-8}$ | Y |
| | | Females | 0.075 | 0.0812 (10,000) | $1.61\times10^{-11}$ | Y |
| | | Males | 0.051 | PRS ns | 0.379 | Y |
| rs12948060 | T | All | 0.062 | 0.0224 (10,000) | 0.411 | Y |
| | | **Females** | **0.156** | **0.0224 (10,000)** | $\mathbf{4.34\times10^{-4}}$ | **Y** |
| | | Males | 0.074 | 0.0017 (10,000) | 0.744 | Y |
| rs35104205 | C | All | 0.059 | 0.0314 (10,000) | 0.571 | Y |
| | | **Females** | **0.154** | **0.0227 (10,000)** | $\mathbf{2.52\times10^{-10}}$ | **Y** |
| | | Males | 0.068 | 0.0021 (10,000) | 0.108 | Y |
| rs4646347 | T | All | 0.062 | 0.0224 (10,000) | 0.412 | Y |
| | | **Females** | **0.156** | **0.0224 (10,000)** | $\mathbf{3.43\times10^{-4}}$ | **Y** |
| | | Males | 0.074 | 0.0017 (10,000) | 0.788 | Y |
| rs4646346 | C | All | 0.062 | 0.0224 (10,000) | 0.383 | Y |
| | | **Females** | **0.156** | **0.0224 (10,000)** | $\mathbf{9.54\times10^{-4}}$ | **Y** |
| | | Males | 0.074 | 0.0017 (10,000) | 0.617 | Y |
| rs7224725 | T | All | 0.023 | 0.0723 (10,000) | $1.63\times10^{-6}$ | N |
| | | Females | 0.036 | 0.164 (1,000) | $1.22\times10^{-7}$ | N |
| | | Males | 0.057 | PRS ns | 0.948 | N |
| rs9944423 | G | All | 0.023 | 0.0625 (10,000) | $1.53\times10^{-6}$ | N |
| | | Females | 0.036 | 0.125 (1,000) | $1.30\times10^{-7}$ | N |
| | | Males | 0.057 | PRS ns | 0.988 | N |

[A] Trait-increasing allele indicates the allele of the *SREBF1* waist-hip-ratio adjusted for body mass index (WHRadjBMI) GWAS variant that results in a positive effect on WHRadjBMI.

[B] Incremental variance explained of WHRadjBMI by the regional WHRadjBMI PRSs built using the *cis* regional variants residing in the adipocyte open chromatin of the *trans* gene sets in the UK Biobank, calculated against a null model containing the covariates of age, age2, the top 20 genetic principal components, testing center, genotyping array (and sex in the all group). The regional WHRadjBMI PRSs of the *trans* gene sets were built excluding the seven *SREBF1* WHRadjBMI GWAS SNPs and their LD proxies (please see Methods).

[C] Significance of each regional PRS was calculated by ranking the observed incremental R2 against the incremental R2 of PRSs similarly built from the *cis* regional variants of random gene sets of the same size as in the actual set. The number of permutations (either 1,000 or 10,000) is listed in parentheses. PRS ns indicates that the regional WHRadjBMI PRS itself did not significantly ($p_{R^2}$>0.05) explain variance of WHRadjBMI, and thus no permutations were performed.

[D] Wilcoxon p-value comparing the regional PRS between the risk allele carriers and non-carriers of the *SREBF1* WHRadjBMI GWAS SNP.

[E] Y indicates that the *trans* gene sets upregulated in the risk alleles carriers of the *SREBF1* WHRadjBMI GWAS variant showed significant pathway enrichment (FDR<0.05) using WebGestalt, while N indicates that no significant enrichments were observed.

Results that are significant in both enrichment of variance explained in WHRadjBMI by the regional PRS and in the Wilcoxon test comparing the magnitudes of the regional PRSs are shown in bold.

**Supplementary Table 18.** Variants residing in the adipocyte open chromatin in the *cis* regions of the downregulated *trans* gene sets reveal significantly enriched ($p_{perm}<0.05$) polygenic risk scores (PRSs) for WHRadjBMI and significant differences in the magnitude of their regional WHRadjBMI PRSs between carriers and non-carriers of the trait-increasing risk allele of the seven *SREBF1* WHRadjBMI GWAS SNPs.

| Variant | Effect allele[A] | Group | $R^2_{WHRadjBMI}$ (%)[B] | Permuted p-value[C] | Wilcoxon p-value[D] | Pathway enriched[E] |
|---|---|---|---|---|---|---|
| rs4924755 | G | **All** | **0.073** | **0.0063 (10,000)** | **$4.47\times10^{-11}$** | **N** |
| | | Females | 0.071 | 0.0636 (10,000) | $1.86\times10^{-88}$ | N |
| | | Males | 0.066 | 0.0011 (10,000) | 0.831 | N |
| rs12948060 | T | All | - | - | - | N |
| | | Females | - | - | - | N |
| | | Males | - | - | - | N |
| rs35104205 | C | All | - | - | - | N |
| | | Females | - | - | - | N |
| | | Males | 0.049 | PRS ns | 0.734 | N |
| rs4646347 | T | All | - | - | - | N |
| | | Females | - | - | - | N |
| | | Males | - | - | - | N |
| rs4646346 | C | All | - | - | - | N |
| | | Females | - | - | - | N |
| | | Males | - | - | - | N |
| rs7224725 | T | All | 0.048 | 0.0096 (10,000) | 0.136 | N |
| | | **Females** | **0.073** | **0.0381 (10,000)** | **$8.03\times10^{-4}$** | **N** |
| | | Males | 0.048 | PRS ns | 0.198 | N |
| rs9944423 | G | All | 0.048 | 0.0096 (10,000) | 0.134 | N |
| | | **Females** | **0.073** | **0.0381 (10,000)** | **$5.65\times10^{-4}$** | **N** |
| | | Males | 0.048 | PRS ns | 0.186 | N |

[A] Trait-increasing allele indicates the allele of the *SREBF1* waist-hip-ratio adjusted for body mass index (WHRadjBMI) GWAS variant that results in a positive effect on WHRadjBMI.

[B] Incremental variance explained of WHRadjBMI by the regional WHRadjBMI PRSs built using the *cis* regional variants residing in the adipocyte open chromatin of the *trans* gene sets in the UK Biobank, calculated against a null model containing the covariates of age, age2, the top 20 genetic principal components, testing center, genotyping array (and sex in the all group). The regional WHRadjBMI PRSs of the *trans* gene sets were built excluding the seven *SREBF1* WHRadjBMI GWAS SNPs and their LD proxies (please see Methods). The regional PRSs were only built in sets with >=5 clumped and thresholded SNPs, and sets with fewer SNPs are indicated with -.

[C] Significance of each regional PRS was calculated by ranking the observed incremental R2 against the incremental R2 of PRSs similarly built from the *cis* regional variants of random gene sets of the same size as in the actual set. The number of permutations (either 1,000 or 10,000) is listed in parentheses. PRS ns indicates that the regional WHRadjBMI PRS itself did not significantly ($p_{R^2}>0.05$) explain variance of WHRadjBMI, and thus no permutations were performed.

[D] Wilcoxon p-value comparing the regional PRS between the risk allele carriers and non-carriers of the *SREBF1* WHRadjBMI GWAS SNP.

[E] Y indicates that the *trans* gene sets upregulated in the risk alleles carriers of the *SREBF1* WHRadjBMI GWAS variant showed significant pathway enrichment (FDR<0.05) using WebGestalt, while N indicates that no significant enrichments were observed.

Results that are significant in both enrichment of variance explained in WHRadjBMI by the regional PRS and in the Wilcoxon test comparing the magnitudes of the regional PRSs are shown in bold.
